# Mepolizumab induced changes in nasal methylome and transcriptome to predict response in asthma

**DOI:** 10.1101/2023.05.18.23290155

**Authors:** Kamini Rakkar, Yik Lam Pang, Poojitha Rajasekar, Michael A Portelli, Robert J Hall, Rachel L Clifford, Dominick Shaw, Ian Sayers

## Abstract

**Rationale:** Mepolizumab is effective for a subset of severe asthma patients in reducing exacerbation frequency. Discovery of a predictive/early marker accurately identifying patients that will have a long-term beneficial clinical response would enable targeting of treatment.

**Objectives:** We aimed to characterise the nasal methylome and transcriptome post Mepolizumab and identify signatures related to responder/non-responder status.

**Methods:** Nasal brushes were taken at baseline (pre-drug) and following 3 months of treatment with Mepolizumab from patients with severe asthma. Both DNA and RNA were extracted. Gene expression was investigated using poly-A RNA sequencing (25M reads) and DNA methylation analysed using the EPIC Array.

**Measurements and Main Results:** 27 paired samples were included, 17 patients were clinical responders and 10 were non-responders at one year. Differential gene expression and DNA methylation analyses identified 6719 genes and 53 CpG sites respectively that changed in response to Mepolizumab. There were 1784 genes which were unique to responders and 893 genes unique to non-responders. Pathway analyses revealed unique gene expression signatures. Respiratory disease associations and regulators of ongoing T2 inflammation pathway were still active in non-responders, and there was an inhibition of neutrophil activation pathways in responders.

**Conclusions:** There was a significant change in both the transcriptome and methylome in the nasal epithelium in patients three months post-Mepolizumab therapy suggesting broad effects on the airway epithelium in severe asthma. Responder and non-responder group analyses indicate there is a responder-specific gene expression profile that may aid in predicting response at one year.

## Introduction

Asthma affects an estimated 300 million people worldwide and was responsible for 21.6 million disability-adjusted life years in 2019 (1). Globally, severe asthma affects between 3-10% of all patients with asthma (2). In severe asthma, symptoms are usually uncontrolled and exacerbation frequency is high, despite the high dose of medication, including inhaled and oral corticosteroids (2). For some, monoclonal antibody therapy is an option and has been used successfully in several large studies, reducing exacerbation frequency and maintenance oral corticosteroid use (3).

Mepolizumab is an injectable anti-IL5 monoclonal antibody, that prevents the binding of IL5 to its surface receptor on eosinophils (4). It reduces exacerbation frequency and steroid burden (5), however the impact on the airway, particularly at the gene level has not been investigated. In addition, typically a third of patients fail to respond to treatment as seen in real-world studies (6). In the UK, current National Institute for Health and Care Excellence (NICE) guidelines determine efficacy after one year of treatment, based on a clinically significant reduction in exacerbations or continuous oral corticosteroid use (TA671 2021). For patients who do not respond, the duration of medication burden is unnecessary and prevents switching to another monoclonal antibody class, of which there are several. It also costs the NHS time and money.

With additional anti-IL5/IL5R therapies (Reslizumab) and Benralizumab and other biologics targeting separate pathways such as Omalizumab (anti-IgE) and Dupilumab (anti-IL4 and IL13) also available, it is important to investigate biomarkers which may be able to predict early responses to a specific drug, and therefore enable better pharmacological management of patients. Furthermore, investigation of molecular mechanisms may uncover why some patients do not respond to treatment and provide greater understanding of asthma.

Transcriptomic analysis of the airway epithelium is proving a useful tool to both understand asthma pathogenesis and the molecular characterization of responders to specific drugs such as inhaled corticosteroids (7). More recently, the nasal epithelium has shown utility in understanding asthma and drug responses (8) and provides an easy but relevant option in investigating molecular mechanisms of the airway epithelium.

We aimed to explore any changes in gene expression and/or DNA methylation in response to Mepolizumab therapy in the nasal epithelium to provide novel drug mechanistic insight. Furthermore, we aimed to detect a predictive or early gene expression and/or DNA methylation signature that could identify responders or non-responders and therefore enable Mepolizumab therapy to be targeted to those most likely to benefit. Asthma has previously been defined with respect to Type 1 (T1) or Type 2 (T2) inflammation, characterised by high neutrophil (T1) or high eosinophil, immunoglobulin E and fractional exhaled nitric oxide (T2) levels. However new data is suggesting a more nuanced approach is required, a statement supported by this study. Our findings suggest there is a significant transcriptional response to Mepolizumab in the nasal epithelium which is, at least in part, mediated by changes in DNA methylation. We also identified a unique gene expression signature in patients that had a clinically significant response at one year.

## Methods

Full methods and statistical analyses are listed in the Supplement. Flow diagram indicating sample and analysis pipeline is in Figure 1.

**Figure 1.**
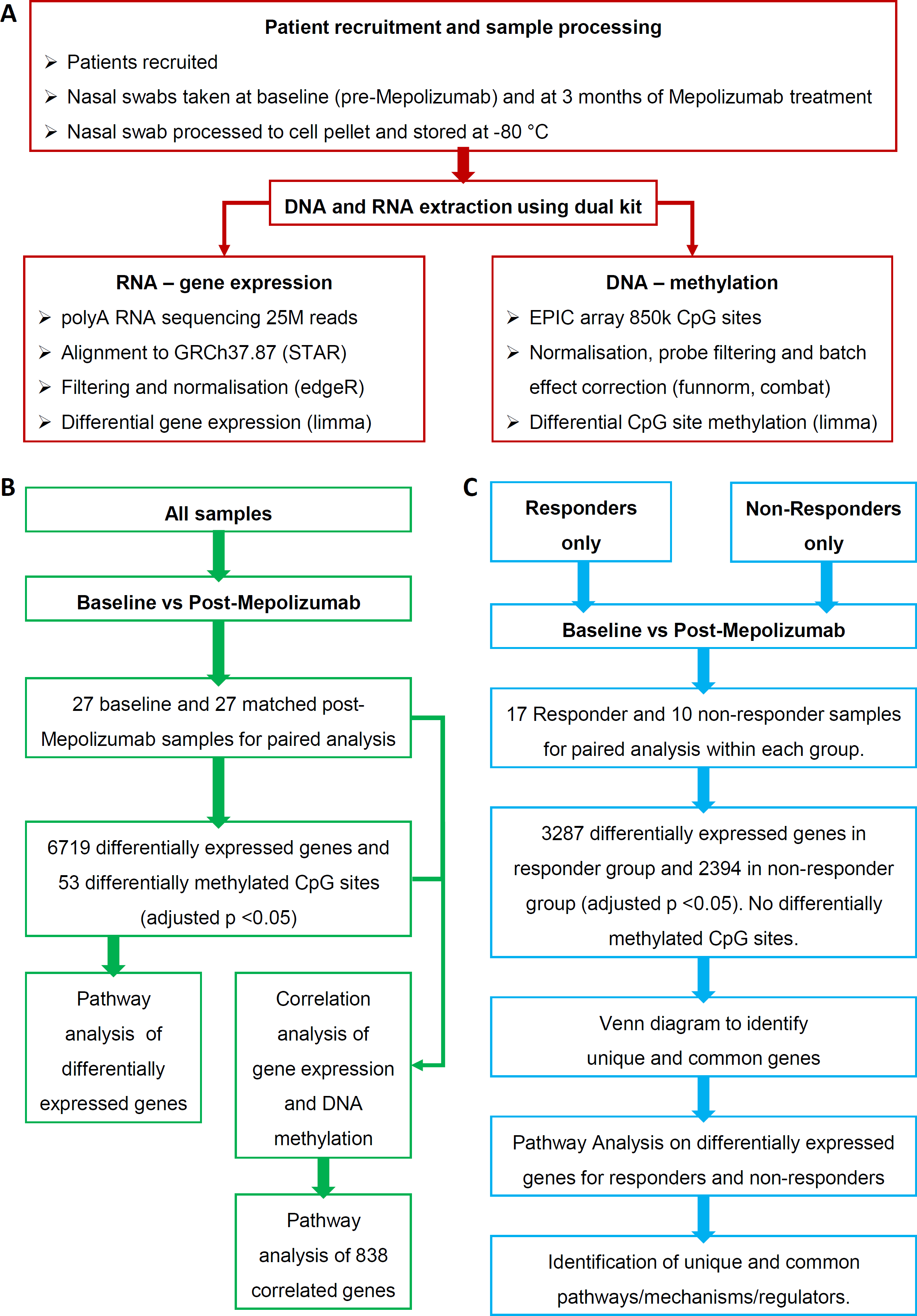
Study design and analysis workflow for gene expression and DNA methylation data. (A) Patients were recruited into the PROCLAIM study and nasal brush samples were taken at baseline and 3 months post-Mepolizumab therapy. DNA and RNA was extracted from the same brush. Gene expression was analysed through polyA RNA sequencing and DNA methylation through the EPIC Array. Gene expression and DNA methylation raw data was evaluated as part of quality control, filtered and normalised before differential analysis. (B) A differential analysis was conducted to compare gene expression and DNA methylation differences between baseline and post-Mepolizumab therapy in all paired samples and a pathway analysis was conducted on the differentially expressed genes. Differentially expressed genes and differentially methylated CpG sites were correlated, and a pathway analysis run on the correlated genes (C) A differential analysis was conducted to compare gene expression and DNA methylation differences between baseline and post-Mepolizumab therapy in either responder or non-responder paired samples alone and pathway analyses were conducted on the differentially expressed genes. An adjusted p value cut off of <0.05 was applied for all data.

### Recruitment

The PROCLAIM study (Poor Response tO monoClonaL therApy In asthMa) enrolled 42 patients with severe asthma who matched the UK National Health Service (NHS) criteria for receiving Mepolizumab (TA431). Patients were recruited between January 2019 and June 2020. The study was approved by the Medical Ethics Committee (REC18/EM/0268). 27 patients’ data was used in this analysis due to patient withdrawal (4 patients) and unavailability of matched nasal brush samples for baseline and 3 months post-Mepolizumab treatment (11 patients).

### Sample collection

Nasal brushes were taken at baseline (pre-drug) and 3-months of Mepolizumab treatment. RNA and DNA were extracted with a dual RNA/DNA Purification (Norgen Biotek Corp.).

### Responder group allocation

Responders were defined as patients who had a 50% reduction in oral corticoid steroid use or 50% reduction in exacerbation frequency after one year of Mepolizumab therapy as per NICE guidelines TA431 (2017). For consistency and adherence to protocol, we did not alter the study’s definition of clinical response when NICE published the new TA671 (2021).

### RNA sequencing

Gene expression was determined through stranded paired end polyA RNA sequencing (Oxford Genomics) using Illumina v1.5 chemistry and the NovaSeq6000 at a minimum of 25M reads per sample.

### DNA methylation

DNA methylation was determined through the Illumina Infinium MethylationEPIC Bead Chip Array (Diagenode) which covers 850,000 methylation sites.

## Results

### Responders and non-responders show no significant differences in clinical or demographic features at baseline

The patient cohort had a median age of 57 years and predominance of females (62%) in line with other severe asthma cohorts (9). As expected, patients had at least one comorbidity (90%) and asthma control questionnaire (ACQ6) scores were in line with severe asthma patients with average scores >3. Similarly, clinical features such as lung function measured by FEV_1_/FVC ratios are <70% and eosinophil levels are 0.55 x10^9^/L or higher, indicative of severe asthma patients (Table 1).

**Table 1.**
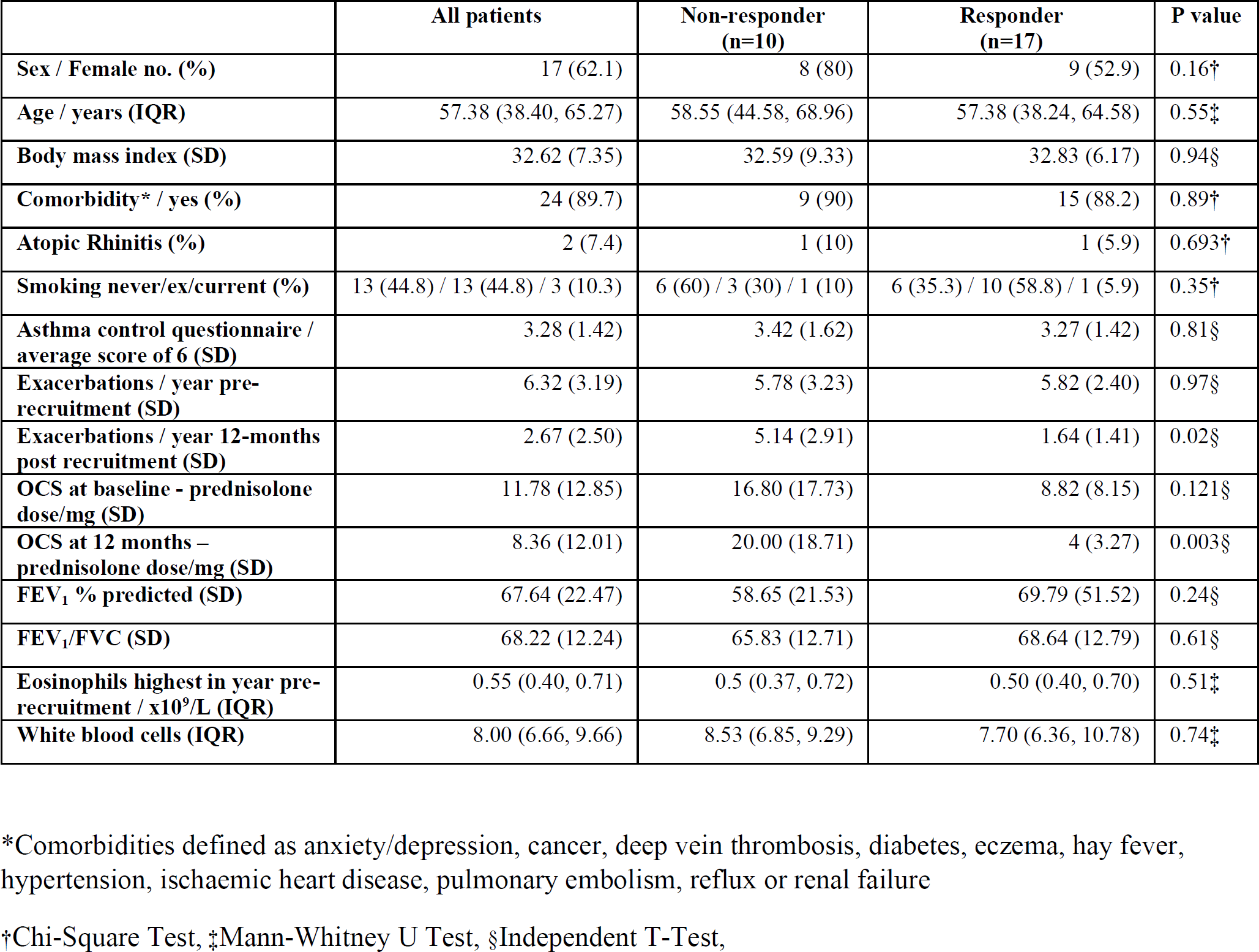
Clinical and demographic features of patients at baseline and/or 12-months (as indicated) and comparison between responder and non-responder groups.

|  | All patients | Non-responder<br>(n=10) | Responder<br>(n=17) | P value |
| --- | --- | --- | --- | --- |
| Sex / Female no. (%) | 17 (62.1) | 8 (80) | 9 (52.9) | 0.16† |
| Age / years (IQR) | 57.38 (38.40, 65.27) | 58.55 (44.58, 68.96) | 57.38 (38.24, 64.58) | 0.55‡ |
| Body mass index (SD) | 32.62 (7.35) | 32.59 (9.33) | 32.83 (6.17) | 0.94§ |
| Comorbidity* / yes (%) | 24 (89.7) | 9 (90) | 15 (88.2) | 0.89† |
| Atopic Rhinitis (%) | 2 (7.4) | 1 (10) | 1 (5.9) | 0.693† |
| Smoking never/ex/current (%) | 13 (44.8) / 13 (44.8) / 3 (10.3) | 6 (60) / 3 (30) / 1 (10) | 6 (35.3) / 10 (58.8) / 1 (5.9) | 0.35† |
| Asthma control questionnaire / average score of 6 (SD) | 3.28 (1.42) | 3.42 (1.62) | 3.27 (1.42) | 0.81§ |
| Exacerbations / year pre-recruitment (SD) | 6.32 (3.19) | 5.78 (3.23) | 5.82 (2.40) | 0.97§ |
| Exacerbations / year 12-months post recruitment (SD) | 2.67 (2.50) | 5.14 (2.91) | 1.64 (1.41) | 0.02§ |
| OCS at baseline - prednisolone dose/mg (SD) | 11.78 (12.85) | 16.80 (17.73) | 8.82 (8.15) | 0.121§ |
| OCS at 12 months – prednisolone dose/mg (SD) | 8.36 (12.01) | 20.00 (18.71) | 4 (3.27) | 0.003§ |
| FEV<sub>1</sub> % predicted (SD) | 67.64 (22.47) | 58.65 (21.53) | 69.79 (51.52) | 0.24§ |
| FEV<sub>1</sub>/FVC (SD) | 68.22 (12.24) | 65.83 (12.71) | 68.64 (12.79) | 0.61§ |
| Eosinophils highest in year pre-recruitment / $\times 10^9/L$ (IQR) | 0.55 (0.40, 0.71) | 0.5 (0.37, 0.72) | 0.50 (0.40, 0.70) | 0.51‡ |
| White blood cells (IQR) | 8.00 (6.66, 9.66) | 8.53 (6.85, 9.29) | 7.70 (6.36, 10.78) | 0.74‡ |

\*Comorbidities defined as anxiety/depression, cancer, deep vein thrombosis, diabetes, eczema, hay fever, hypertension, ischaemic heart disease, pulmonary embolism, reflux or renal failure
†Chi-Square Test, ‡Mann-Whitney U Test, §Independent T-Test,

### The nasal cell composition following Mepolizumab treatment is unaltered

After filtering and normalisation 684,807 probes were used in the methylation analysis (Supplement Figure 1). A reference dataset (EpiDISH) was used to determine indirectly the cell type composition of the nasal brush samples using DNA methylation data. Epithelial cell, fibroblast, and immune cell markers were analysed. There were no significant differences in inferred cell type composition between baseline and post-Mepolizumab therapy and between responder and non-responder groups (Supplement Figure 2). Epithelial cells were the largest observed population of cells.

### Mepolizumab induces significant changes in gene expression in the nasal epithelium

Principal component analysis of gene expression shows distinct clustering of patients post-Mepolizumab therapy (red dots) compared to a more heterogenous distribution pre-treatment (blue dots, Figure 2A). A total of 2547 genes were down-regulated, and 4172 genes were up-regulated following 3 months treatment in all patients (Figure 2B). Expression values of these genes in each patient sample distinctly shows the changes in expression after therapy (Figure 2C). A list of differentially expressed genes is provided (Supplement Table 1).

**Figure 2.**
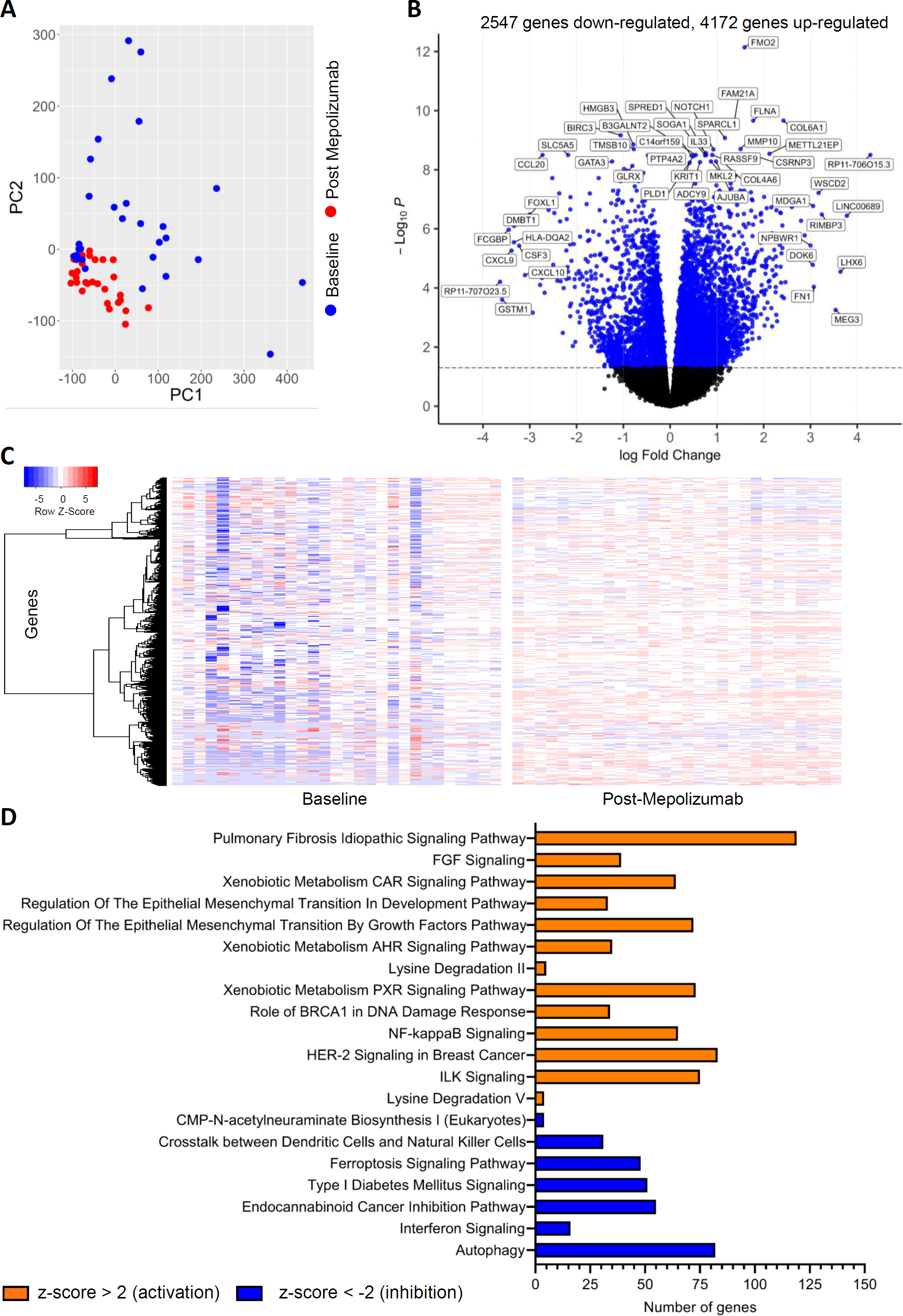
Mepolizumab induces gene expression changes in the nasal epithelium of severe asthma patients. (A) Principal component analysis shows clustering of patients’ gene expression after Mepolizumab therapy. (B) Volcano plot of baseline vs post-Mepolizumab differential gene expression in all subjects. (C) Heatmap of the differentially expressed genes grouped into baseline and post-Mepolizumab blocks. (D) Pathways associated with Mepolizumab-induced gene expression changes. An adjusted p value of <0.05 was applied for all data.

### Mepolizumab-induced changes in gene expression are associated with downstream viral disease

Disease association pathway analysis of the gene expression changes following Mepolizumab therapy, identified significant associations with multiple viral related pathways e.g. viral infection, release of lentivirus. Other significant associations were related to inflammatory diseases such as psoriasis and immune related inflammatory disease. A full list of disease associations is provided (Supplement Table 2).

### Mepolizumab-induced changes in gene expression are associated with inflammation, xenobiotic metabolism and fibrosis pathways

Pathway analysis of the gene expression changes following Mepolizumab therapy show evidence of suppression of inflammatory pathways. For example, pathways related to crosstalk between dendritic cells and natural killer cells, “type 1 diabetes and interferon signalling were observed. There is also evidence of activation of fibrotic pathways e.g. FGF signalling, epithelial mesenchymal transition and pulmonary fibrosis idiopathic signalling pathways (Figure 2D). Full pathway analysis is provided (Supplement Table 3).

### Mepolizumab-induced changes in gene expression are associated with upstream cytokine activity

Upstream analysis is used to identify regulators that may be responsible for the observed gene expression changes. Mepolizumab-induced changes in gene expression were identified to, at least in part, be due to activity changes in asthma related cytokines such as IL13, IFN-alpha, IL1-beta and TNF-alpha (Figure 3A, orange circles) and transcription regulators, for example SP1 and STAT1 (blue circles). Full upstream regulator analysis is provided (Supplement Table 4).

**Figure 3.**
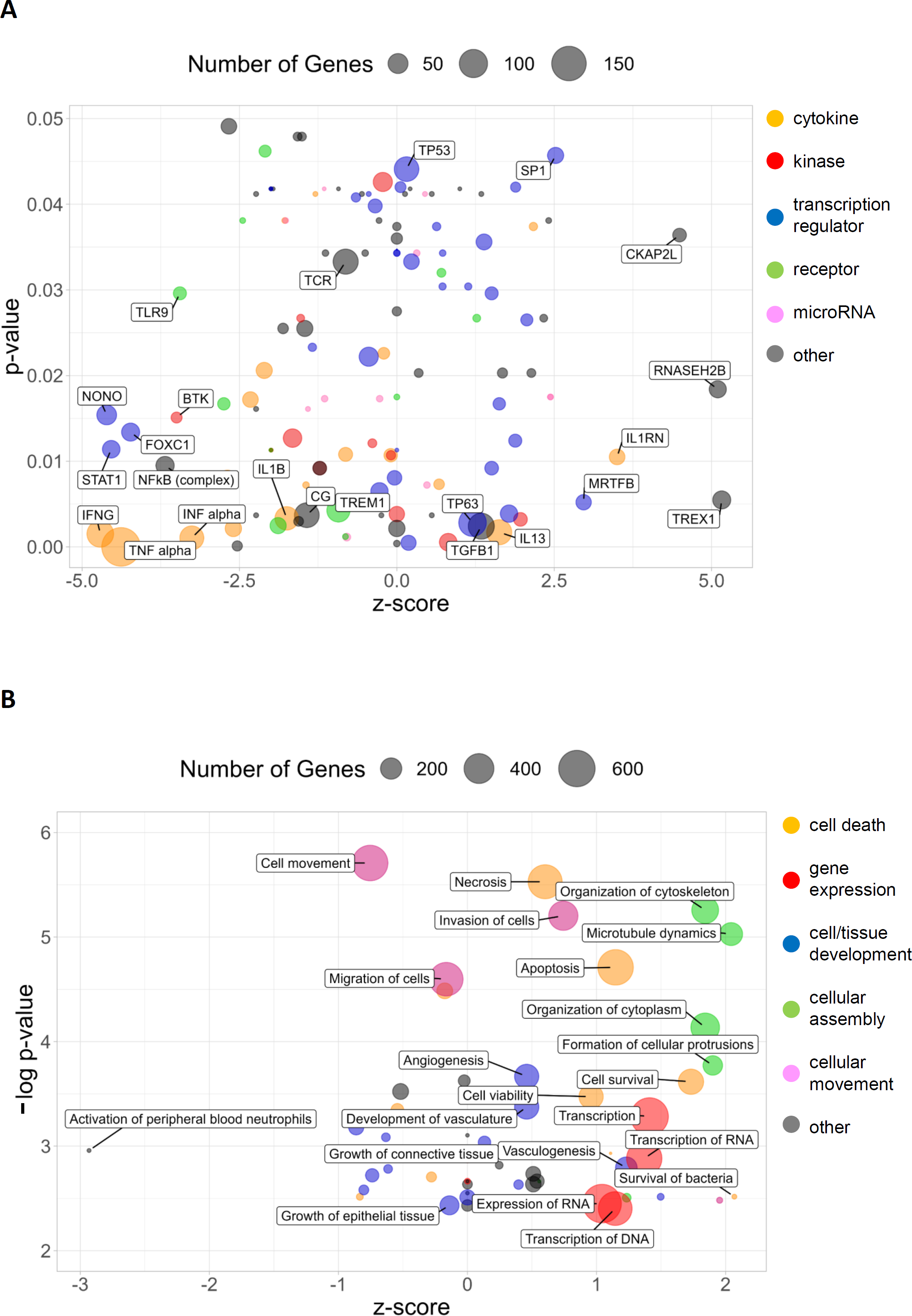
Mepolizumab-induced gene expression changes are associated with cytokine activity. (A) A bubble plot of upstream regulator associations with Mepolizumab-induced gene expression. Upstream regulators are plotted according to p-value and z-score, with the size of the bubble indicating the number of genes in the dataset affected by the regulator. (B) A bubble plot of downstream functional pathway associations with Mepolizumab-induced gene expression. Pathways are plotted according to -log(p-value), due to the broad range of p-values from the data, and z-score, with the size of the bubble indicating the number of genes in the dataset affected by the pathway. An adjusted p value of <0.05 was applied for all data.

### Mepolizumab-induced changes in gene expression are associated with a decrease in downstream neutrophil activation

Downstream functional effects analysis is used to identify functional pathways expected to be activated or inhibited, given the observed expression in the dataset. Mepolizumab-induced gene expression changes show a significant association with an inhibition of activation of peripheral blood neutrophils pathways (z-score of -2.9, Figure 3B). A general activation in gene expression (Figure 3B, red circles) and cellular assembly pathways (green circles) is also observed. Full downstream functional effects analysis is provided (Supplement Table 5).

### There are significant differences in gene expression between responders and non-responders after Mepolizumab therapy

Differential analysis between baseline and post-Mepolizumab treatment gene expression in the responder group alone resulted in 1142 down regulated and 2145 up-regulated genes (adjusted p-value <0.05) (Figure 4A), while 1012 down-regulated and 1382 up-regulated genes were identified in the non-responder group alone (adjusted p-value <0.05) (Figure 4B). Of these, 1784 genes were unique to responders alone and 893 genes were unique to non-responders alone, with the remaining 1500 genes (36%) overlapping (Figure 4C). However, some caution should be used in interpretation due to the differences in responder (n=17)/non-responder (n=10) group sample size. At baseline we found no significant differences in gene expression between responders and non-responders. A list of unique differentially expressed genes for the separate responder or non-responder groups is provided (Supplement Tables 6 and 7).

**Figure 4.**
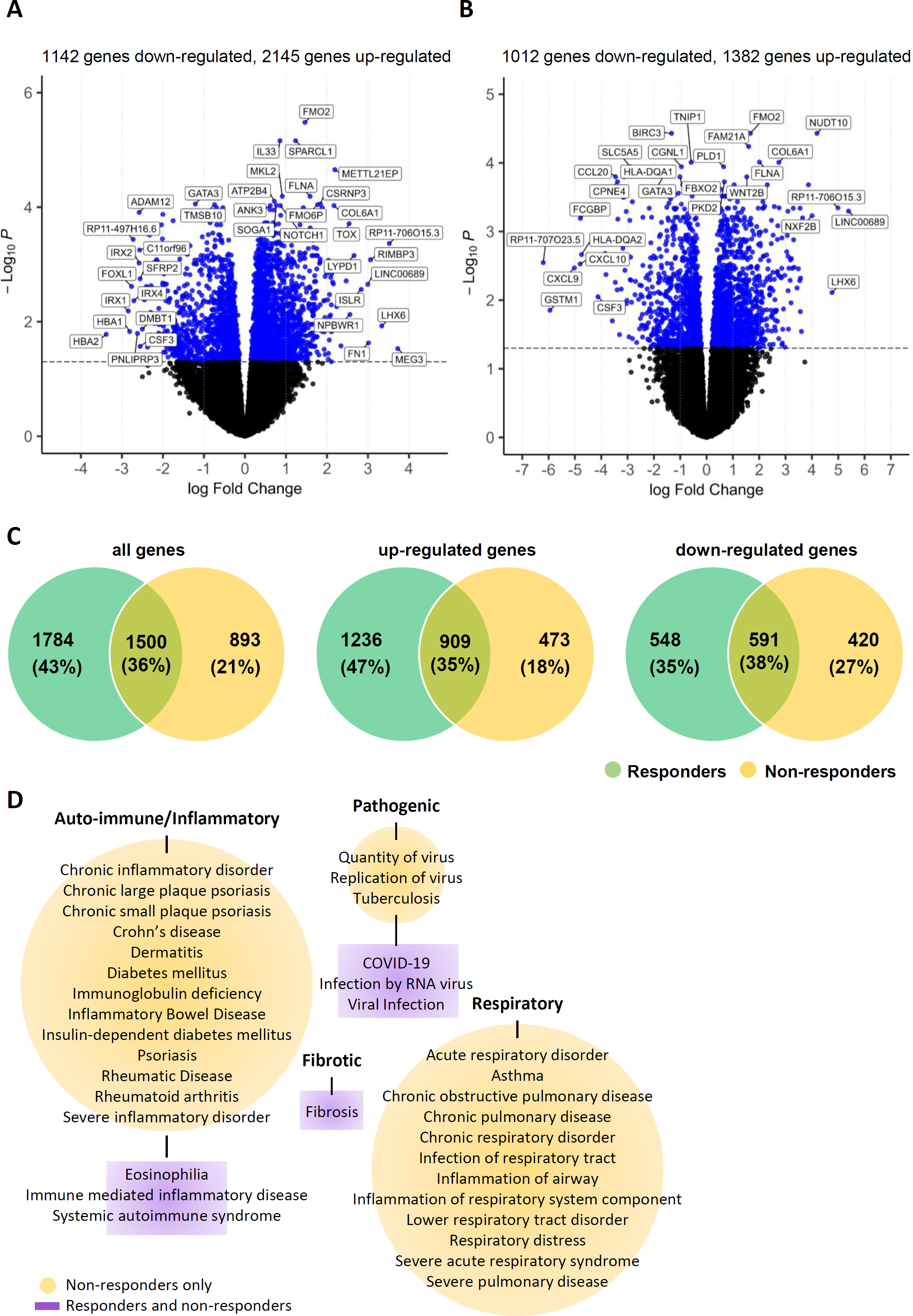
There are differences in gene expression between responders and non-responders at one year following 3 months of Mepolizumab treatment. (A) Volcano plot of baseline vs post-Mepolizumab differential gene expression in responders alone. (B) Volcano plot of baseline vs post-Mepolizumab differential gene expression in non-responders alone. (C) Venn diagram to show overlap of differentially expressed genes between responder and non-responder groups. (D) Disease associations from pathways analysis in separate responder and non-responder groups. Diseases have been grouped into type. Diseases circled in yellow indicate associations common to both responder and non-responder groups. Purple rectangles indicate associations found in the non-responder group only. Due to a large number of disease associations only auto-immune, infectious, inflammatory, or respiratory diseases have been shown. There were no associations found in the responder group alone. An adjusted p value of <0.05 was applied for all data.

### Mepolizuma b-induced gene expression changes are associated with downstream respiratory disease in non-responders but not responders

Disease association pathway analysis of separate responder and non-responder Mepolizumab-induced changes in gene expression, commonly tagged broad terms such as fibrosis, viral infection, immune mediated inflammatory disease and eosinophilia. A large group of specific respiratory and auto-immune disease associations were only present in the non-responder group (Figure 4D). Full disease analysis is provided (Supplement Table 8).

### Mepolizumab-induced changes in gene expression are associated with upstream cytokine activity in non-responders

Upstream regulator analysis in separate responder and non-responder groups showed an overlap of 170 regulators (adjusted p-value <0.05). Mepolizumab-induced gene expression changes were associated with an activation of upstream regulators of TSLP, IL4 and IL33 cytokines in the non-responder dataset only (Figure 5A, orange circles) suggesting there is ongoing T2 inflammation in the airways. Full upstream regulator analysis is provided (Supplement Table 9).

**Figure 5.**
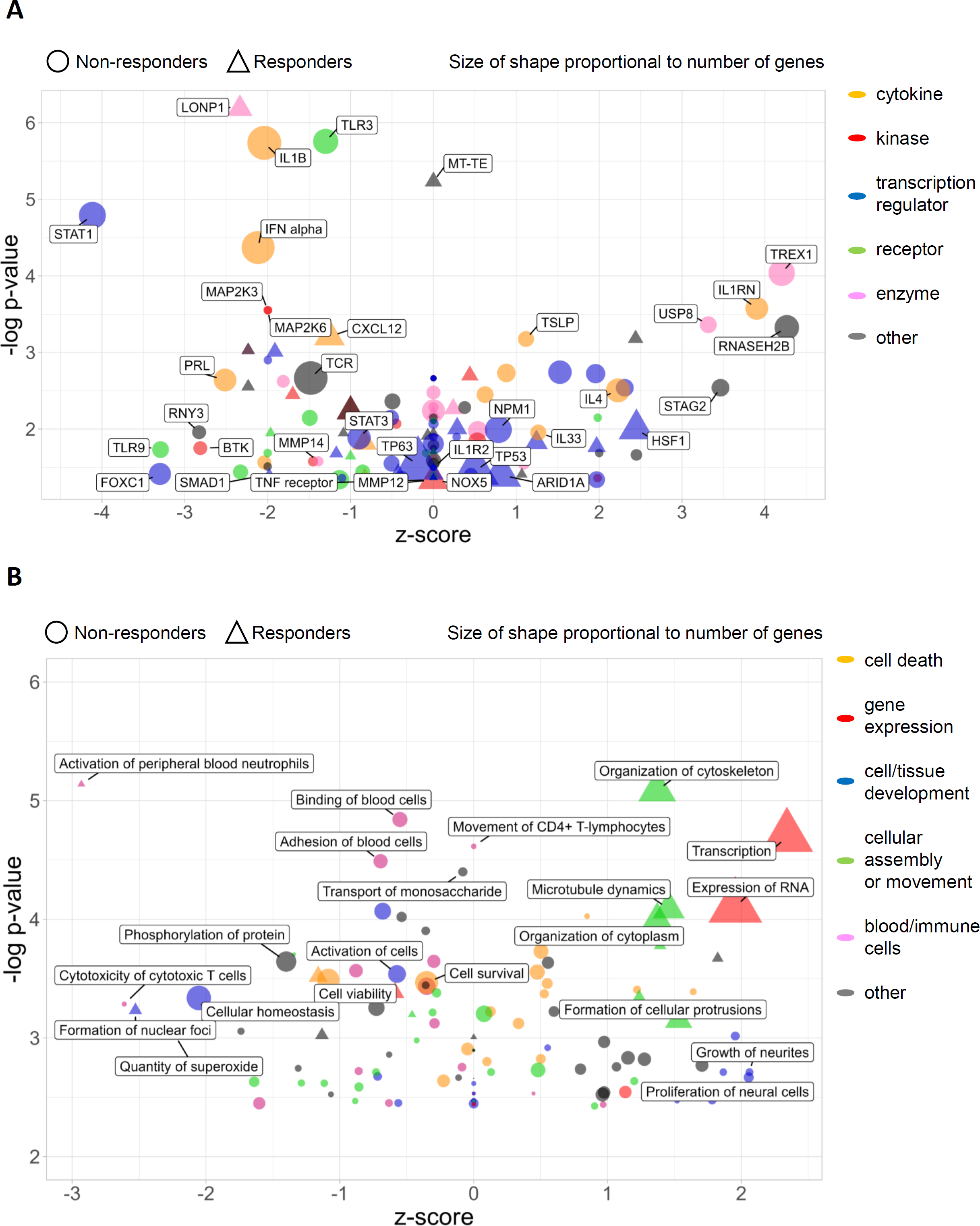
Specific differentially expressed genes post-Mepolizumab therapy are associated with an increase in upstream cytokine regulators in non-responders and an increase in intracellular assembly pathways in responders. (A) A bubble plot of upstream regulator associations which were unique to responders or non-responders (associations common to both groups not shown). Upstream regulators are plotted according to -log(p-value), due to the broad range of p-values from the data, and z-score, with the size of the bubble indicating the number of genes in the dataset affected by the regulator. (B) A bubble plot of downstream functional pathway associations which were unique to responders or non-responders (associations common to both groups not shown). Pathways are plotted according to -log(p-value), due to the broad range of p-values from the data, and z-score, with the size of the bubble indicating the number of genes in the dataset affected by the pathway. An adjusted p value of <0.05 was applied for all data.

### Mepolizumab-induced changes in gene expression are associated with a decrease in downstream neutrophil activation in responders but not non-responders

Mepolizumab induced gene expression changes drove different downstream pathways in responder and non-responder datasets. A responder signature included suppression of pathways related to activation of peripheral blood neutrophils, quantity of superoxide, apoptosis and cell death of epithelial cells and increases pathways related to intracellular assembly pathways including; microtubule dynamics, organization of cytoplasm and cytoskeleton (Figure 5B, triangles). In the non-responder dataset, there was an inhibition in pathways related to cellular homeostasis, adhesion of immune cells, cell survival viability. Pathways related to cell death of immune cells and fibroblast development were modestly activated (Figure 5B, circles). Full downstream functional effects analysis is provided (Supplement Table 10).

### Mepolizumab treatment induces changes in DNA methylation in the nasal epithelium

Principal component analysis (Figure 6A) of DNA methylation did not show any distinct clustering of patients post-Mepolizumab therapy (red dots) compared to baseline (blue dots) in contrast to the gene expression data. A total of 53 CpG sites were differentially methylated (FDR p<0.05) after 3 months of Mepolizumab treatment (Figure 6B). Methylation values of these sites in each patient sample show the changes in DNA methylation after therapy (Figure 6C). Full list of differentially methylated CpG sites is provided (Supplement Table 11).

**Figure 6.**
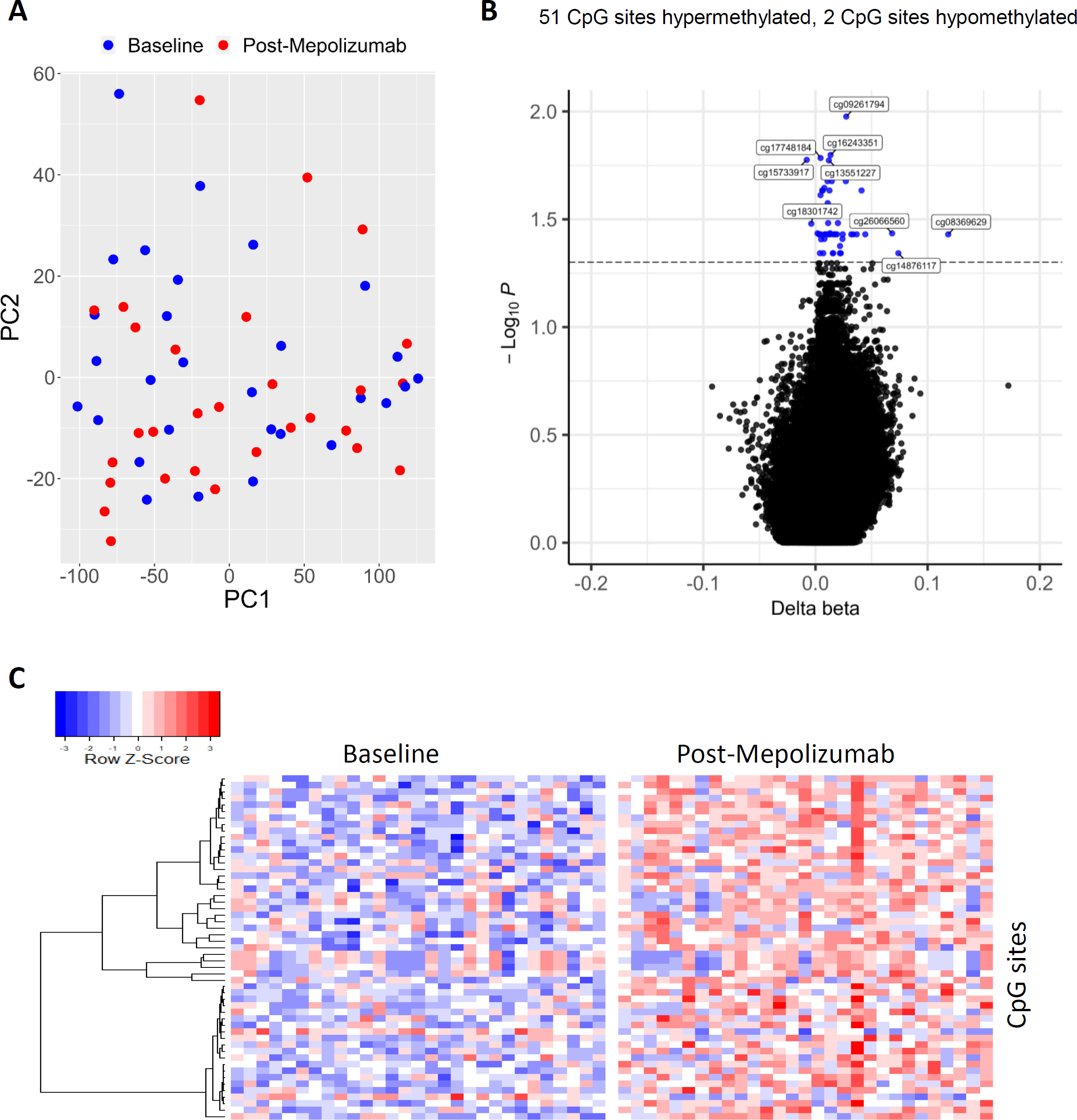
Mepolizumab induces DNA methylation changes in the nasal epithelium of severe asthma patients. (A) Principal component analysis shows no clustering of patients’ DNA methylation profiles after Mepolizumab therapy. (B) Volcano plot of baseline vs post-Mepolizumab differential DNA methylation. (C) Heatmap of the differentially methylated CpG sites grouped into baseline and post-Mepolizumab blocks. An adjusted p value of <0.05 was applied for all data.

### Genes which correlate with Mepolizumab-induced changes in DNA methylation are associated with primary ciliary dyskinesia, cellular assembly, movement and metabolism

There were 1452 significant DNA methylation and gene expression correlations. This involved 42 (79%) of the differential methylated sites and 838 (12%) of differentially expressed genes (Figure 7A and B). Disease pathway analysis showed a strong association with respiratory diseases such as primary ciliary dyskinesia, chronic familial respiratory disorder and sinus disorder (Figure 7C). Full correlation and disease analyses is provided (Supplement Tables 12 and 13).

**Figure 7.**
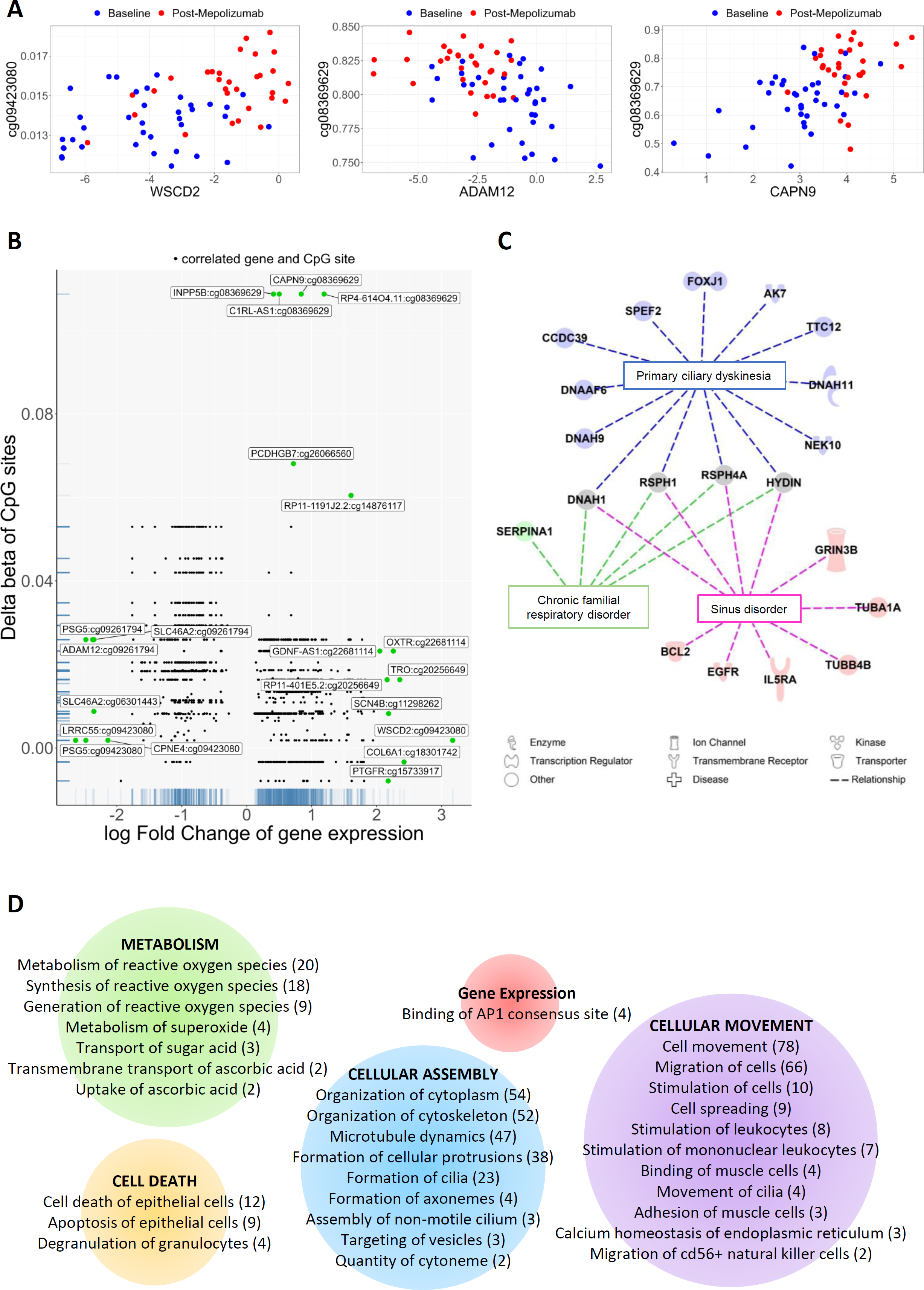
Mepolizumab induced changes in gene expression and DNA methylation correlate with each other. (A) Representative correlations between 3 different genes and correlated CpG sites. Blue dots show samples at baseline and red dots show samples post-Mepolizumab therapy. (B) Plot of all CpG site and gene correlations. Each dot represents a correlation between and CpG site and gene. (C) Diseases associated with 838 differentially expressed genes which were correlated with 42 of the differentially methylated CpG. (D) Downstream functional pathways associated with these 838 genes. Pathways have been grouped into types and the number of genes in each pathway are in brackets. An adjusted p value of <0.05 was applied for all data.

Downstream function analysis of correlated genes show association with pathways predominantly involved in cell movement, migration of cells, organisation of the cytoplasm and metabolism of reactive oxygen species (Figure 7D). Full downstream functional effects analysis is provided (Supplement Table 14).

No significant changes in DNA methylation were seen when comparing responder and non-responder methylation profiles at baseline or when comparing baseline and post-Mepolizumab therapy DNA methylation in responder or non-responder datasets separately.

## Discussion

This study set out to identify changes in gene expression and/or DNA methylation in response to Mepolizumab in the nasal epithelium in patients with severe asthma to provide novel understanding of drug mechanisms, and to test the hypothesis that there may be an identifiable gene signature present at three months that predicts clinical response at one year. Overall, these data suggest that 3 months of Mepolizumab treatment significantly impacts airway epithelial gene expression and, by inference, homeostasis in severe asthma, leading to a potentially more homogeneous profile than at recruitment. Of note, there was a suppression of inflammation pathways, particularly those related to viruses/interferons. Interestingly, when gene expression analyses were completed separately in responders/non-responders, distinct gene expression profiles were identified. Non-responders exhibited, amongst others, elevated upstream activation of IL33, TSLP and IL4, suggesting ongoing T2 inflammation remains following treatment, furthermore, respiratory disease associations also remain. In contrast, in responders, gene expression associated with a decrease in downstream neutrophil activation was a feature. Finally, we also investigated the effect of Mepolizumab on DNA methylation and illustrate that a large proportion of differential methylation post treatment is associated with gene expression differences demonstrating that methylation at least in part contributes to the mechanisms driving altered gene expression. In post-Mepolizumab samples *TET3* also showed a slight but significant increase in expression (logFC 0.25) indicating Mepolizumab may be altering DNA methylation mechanisms. These novel data provide a unique insight into the impact of Mepolizumab in the airway epithelium and highlight potential signatures found in the clinical responder population.

To our knowledge, this is the first study to investigate gene expression and DNA methylation in the nasal epithelium of severe adult asthma patients in response to Mepolizumab treatment. Mepolizumab has previously been shown to induce elevated tight junction formation and cilium organisational genes in nasal scrapings from aspirin exacerbated respiratory disease patients (10), which we have also observed in our own results, especially those correlated with DNA methylation. In this study, we show that Mepolizumab has a broad effect on nasal epithelial gene expression, providing new insight and potentially confirming the hypothesis that there is an anti-inflammatory effect on the airway epithelium. This anti-inflammatory property is driven not only by the observation that several chemokine genes were supressed, including CXCL5, 10 and 11, all known to be important in asthma, but also through more novel findings, e.g. changes in pathways related to fibrosis.

When looking at Mepolizumab-induced gene changes in all asthma patients, cytokines such as IL13, and IL1RN are activated in pathways predicted to explain these gene expression changes, whereas IFN-alpha and -gamma, IL1-alpha and -beta, IL17A, and TNF-alpha are predicted to be inhibited. Other studies have also shown cytokines to be affected by Mepolizumab therapy. One study found IL1 receptor antagonist to be increased after 4 weeks of Mepolizumab therapy (11) whereas in another study serum levels of TSLP were reduced after Mepolizumab therapy (12).

The current study has taken a molecular approach in investigating a response signature. Other studies have focussed on genetic associations (13), blood (14) or sputum biomarkers (15), clinical features such as lung function (16) or even characterising super responders (17) but have found limited evidence. However, ACQ scores have been shown to be predictors of response (18).

Explorative analysis into specific responder or non-responder groups alone, shows that, IL4, IL33, and TSLP are activated and IL1-alpha and -beta and IFN-alpha 2 are inhibited in non-responders only. These results indicate that overall non-responders are experiencing a higher degree of inflammation that is both epithelial and inflammatory cell driven. Furthermore, inhibition of IL-1 and IFN activities could indicate inadequate responses to infections and therefore poor outcome (19). This complexity of the cytokine environment requires further investigation and may in part explain differences in responses and may advocate for alternative methods of treatment. Therapies aimed at targeting the upstream epithelial alarmins such as TSLP (Tezepelumab) and IL33 (Itepekimab) may provide an alternative or dual therapy for patients who do not respond to Mepolizumab (20).

Another gene in our study, CCL5, was found to be unique to responders and had a slight decrease in activity (logFC -0.56). This is of particular note as it is a chemoattractant for eosinophils (21). Another chemoattractant CCL4 has been shown to be lower in the blood of responders compared to non-responders at baseline and after 4 weeks of Mepolizumab therapy (22).

As well as cytokines, investigation of alternate immune cells such as neutrophils may also benefit those patients who do not respond to Mepolizumab. Although we did not see a change in neutrophil numbers in the blood post-treatment (data not shown), at the molecular level, Mepolizumab-induced changes in gene expression were associated with a downstream suppression of the neutrophil activation pathway only in patients who responded to treatment. This may provide an additional mechanism of action of Mepolizumab. In a recent study in asthma patients challenged with rhinovirus, Mepolizumab prevented an increase neutrophil numbers and activation in sputum (23). Similarly, in a study of nasal lavage gene expression, a module of genes related to neutrophil chemotaxis, was found to be inversely associated with exacerbations (24).

In the same nasal lavage study, increased expression of epithelium related gene modules was observed after 52 weeks of Mepolizumab therapy and associated with exacerbation (24). Structural, cellular and tissue changes, such as fibrotic pathways, were also associated with Mepolizumab induced gene expression changes in our study, indicating that Mepolizumab may affect cellular structure and possibly airway remodelling. Although there are no current studies that have directly investigated the role of Mepolizumab in airway remodelling, a study looking at Lebrikizumab, which neutralises IL13, suggests it may reduce asthmatic airway remodelling (25). Interestingly these structural cell biology mechanisms are also strongly correlated with changes in DNA methylation. Previous studies have shown differentially methylated regions to be associated with asthma and relevant genes such as IL4 and IL13 (26). DNA methylation changes have also been associated with cytoskeletal remodelling in asthma (27). Collectively these data and our study suggest cellular structural changes and airway remodelling is a key mechanism that needs to be explored further.

This is the first time the effect of Mepolizumab on gene expression and DNA methylation in the nasal epithelium of patients with severe asthma has been studied. We have found that Mepolizumab affects the inflammatory profile, neutrophil activation, and cellular structural mechanisms in the upper airway epithelium. From our explorative responder/non-responder analysis, our data suggests that there may be an early gene expression signature to indicate response, predominantly related to a reduction in the T2 inflammatory profile in responders but a sustained T2 inflammatory profile in non-responders. However, due to the differences in sample size and therefore statistical power between the responder and non-responder groups, these results need to be interpreted with caution and replication would be of benefit to add confidence to the results but to our knowledge there is no replication cohort with nasal brush samples for which this can be done at the moment. Finally, we have found that changes in DNA methylation associate with gene expression and these are related to cellular assembly, metabolism and movement. Taken together, our data has provided novel molecular insight into the effect of Mepolizumab therapy in severe asthmatic patients and provides a basis for further investigation into molecular mechanisms of response that may lead to a more tailored approach to prescribing.

## Data sharing

The RNA sequencing and DNA methylation data will be deposited in a publicly available database upon acceptance for publication. Summary data is available in the supplements.

## Data Availability

All data produced in the present study are available upon reasonable request to the authors.

## Acknowledgements

Mohammed Ali for his help in administration, recruitment and sample processing. Laura Matthews, Helen Bailey, Carly Clayton, Karina Bingham, and Rebecca M Cooper for their help in sample acquisition and processing.

## Contributions

IS, DS and YLP designed the study. KR processed samples, extracted the RNA and DNA and analysed the data. PR and RLC co-analysed the DNA methylation data. MAP and RH co-analysed the RNA sequencing data. All authors contributed to the writing of the manuscript.

## Supplement

## Methods

### Recruitment

The PROCLAIM study (Poor Response tO monoClonaL therApy In asthMa) recruited 42 patients with severe asthma who matched the National Health Service (NHS) criteria for receiving Mepolizumab (TA431) between January 2019 and June 2020 (1). The study was approved by the Medical Ethics Committee (REC18/EM/0268).

#### Inclusion criteria

All patients will be able to give informed consent for study participation

All patients identified as being suitable for anti-IL5 therapy by the regional severe asthma multidisciplinary team (MDT) at Nottingham Universities Hospitals NHS Trust was considered for study enrolment. This assessment by the MDT for participants to be eligible includes:

- Confirmation of asthma diagnosis
- Historic blood eosinophil counts,
- Annual asthma exacerbation rates
- Oral Prednisolone dose.
- Patients must demonstrate over 75% compliance with inhaled corticosteroid therapy
- The asthma MDT and difficult asthma clinic normally assess patients between 18-80 years so only patients in this age range will be approached
- Confirmation of treatment with anti-IL5 requires at least two asthma specialists to agree that this is the correct course of action

#### Exclusion criteria

Pregnancy

### Sample collection

Nasal brushes were taken at baseline (pre-drug) and 3-months of Mepolizumab treatment and were immediately placed in 3 mL of cold PneumaCult-Ex cell culture media (Stemcell) and stored on ice until processing. Nasal brushes were vortexed and scraped to dislodge cells into the media. The brush was discarded, and the cell suspension centrifuged at 200g. The media supernatant was removed, and cell pellet stored at -80 °C until dual RNA/DNA Purification (Norgen Biotek Corp.). DNA was stored at -20 °C and RNA was stored at -80 °C until use.

### Responder group allocation

Responders were defined as patients who had a 50% reduction in oral corticoid steroid use or 50% reduction in exacerbation frequency after one year of Mepolizumab therapy.

### RNA sequencing

Gene expression was determined through stranded paired end polyA RNA sequencing (Oxford Genomics) using Illumina v1.5 chemistry and the NovaSeq6000 at a minimum of 25M reads. Raw data was aligned to the GRCh37 human assembly using STAR. All further data processing and analysis was undertaken with different packages in RStudio (2). Raw read counts were transformed to counts per million values, unexpressed transcripts removed, and the data normalised using edgeR (v3.38.1). Paired sample weighted differential gene expression, with epithelial cell proportions as calculated below, was determined using Limma (v3.52.1) (3).

### DNA methylation

DNA methylation was determined through the Illumina Infinium MethylationEPIC Bead Chip Array (Diagenode) which covers 850,000 methylation sites. Raw data was processed and analysed using packages in RStudio. Data was normalised using FunNorm (minfi v1.42.0). Probes with less than 3 beads, low detection p values, SNPs at CpG target sites and single base extension sites for type I probes, and which were cross reactive and XY chromosomes specific were removed (Figure 1) (4). Data was batch effect corrected for array and array position using ComBat (sva v3.44.0). The proportion of epithelial cells in each sample was determined using EpiDISH (v2.12.0) (5) and differential CpG site methylation was determined using Limma (v3.52.1) (6).

### Gene expression and DNA methylation correlation

All differentially expressed genes and CpG sites from the baseline vs post-Mepolizumab analysis were correlated in RStudio using Limma (v3.52.1).

### Pathway Analysis of gene expression

The Ingenuity Pathway Analysis tool (Qiagen) was used to determine disease, pathway, upstream regulator, and downstream functional effect associations for identified gene lists. Whole gene expression datasets with adjusted p-values and log fold changes in gene expression were used. Where there was enough data available IPA also calculated Z-scores indicating whether the particular pathway had an increase (positive z-score) or decrease (negative z-score) in activity. For the correlated DNA methylation and gene expression dataset a gene list without numerical data was used as these correlations were not specific to a timepoint and therefore the matched gene expression data could not be used, consequently Z-scores and directionality of pathway associations were not determined. An adjusted p value of <0.05 was applied for all associations and the whole dataset itself was used as the reference (7).

### Statistical analyses

Patient clinical and demographic data were analysed in SPSS (v28). Data were tested for normality using the Shapiro-Wilk test. Those with a normal distribution were analysed using the Independent Student’s T-test. Data with non-normal distributions were analysed using a Mann Whitney U test. Categorical data were analysed using the Chi-Square Test of Independence. P values of <0.05 were considered significant. For DNA methylation, data was analysed using linear modelling and empirical Bayes. For RNA sequencing, data was analysed using precision weighted linear modelling and empirical Bayes. For the DNA methylation and gene expression correlation a linear model was used. Adjusted p values (Benjamini-Hochberg correction) of <0.05 were considered significant.

## Results

**Supplementary Figure 1.**
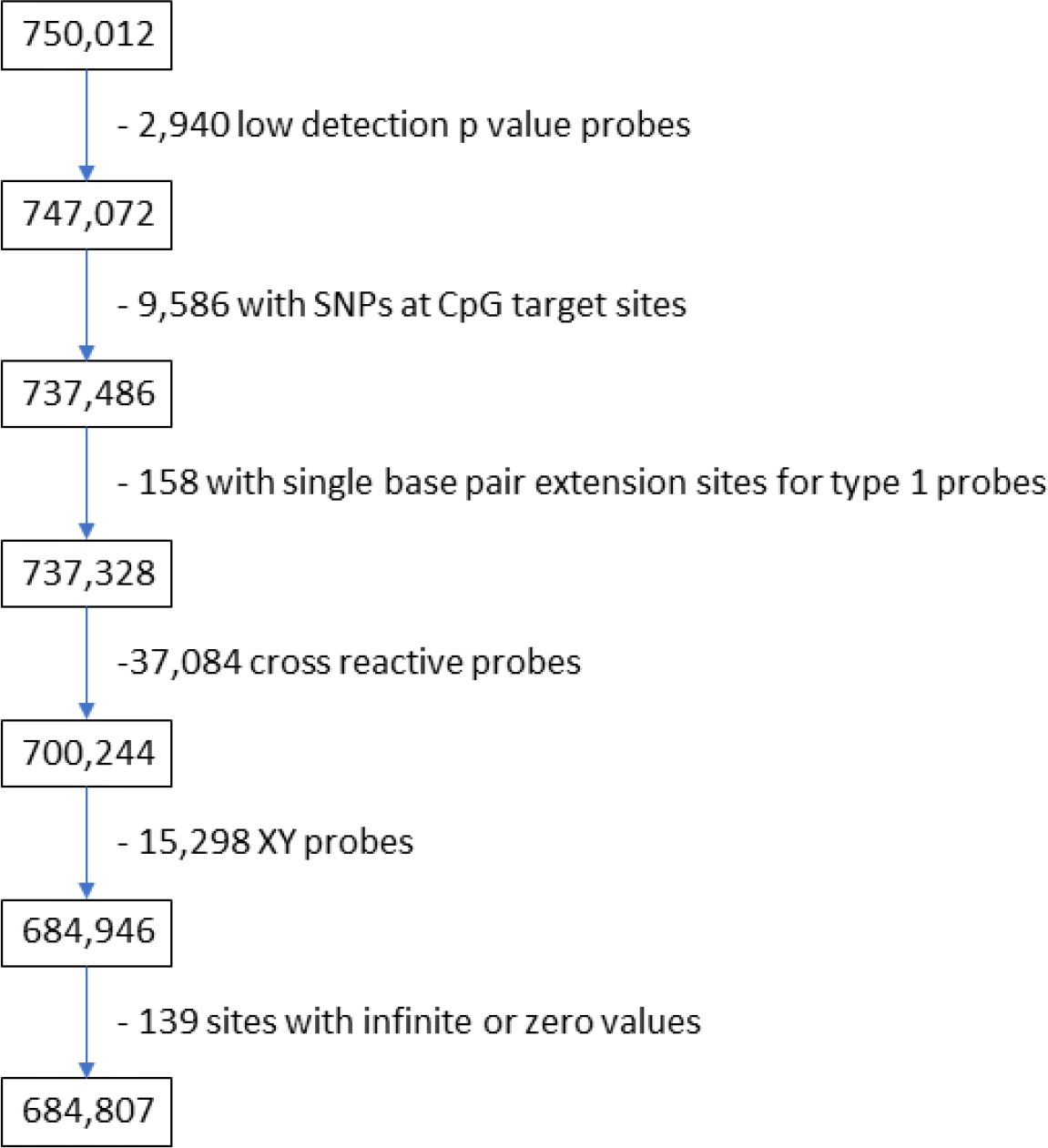
Number of probes removed at each stage of QC for the DNA methylation analysis. After all QC steps 684,807 probes remained in the analysis.

**Supplementary Figure 2.**
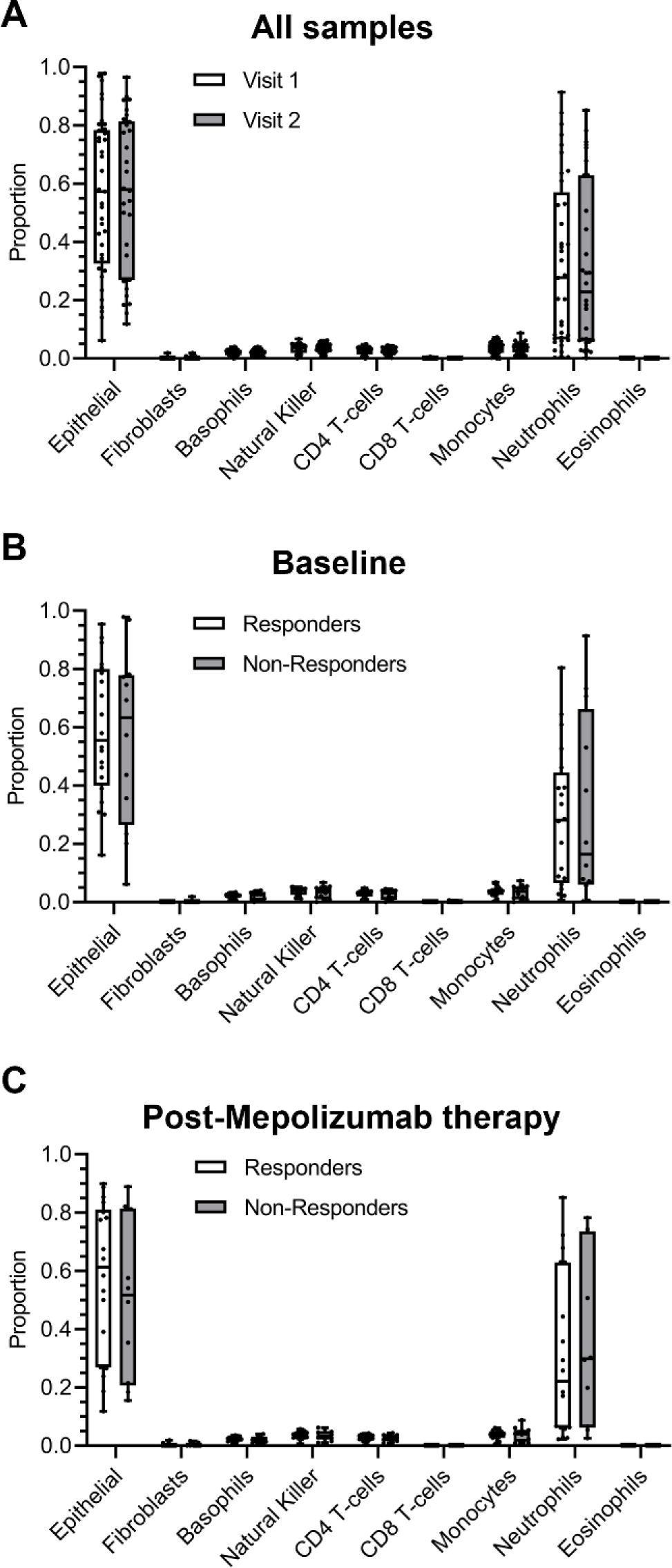
Cell type composition based on DNA methylation data. The EpiDISH package in R was used to determine the cell type composition of the nasal brush samples. Plots show proportion of each cell type for each sample in (A) all samples, and samples at (B) baseline and (C) post-Mepolizumab therapy stratified by visit or responder status. The box and whisker plots show the minimum, first quartile, median, third quartile, and maximum proportional values.

**Supplementary Table 1.**
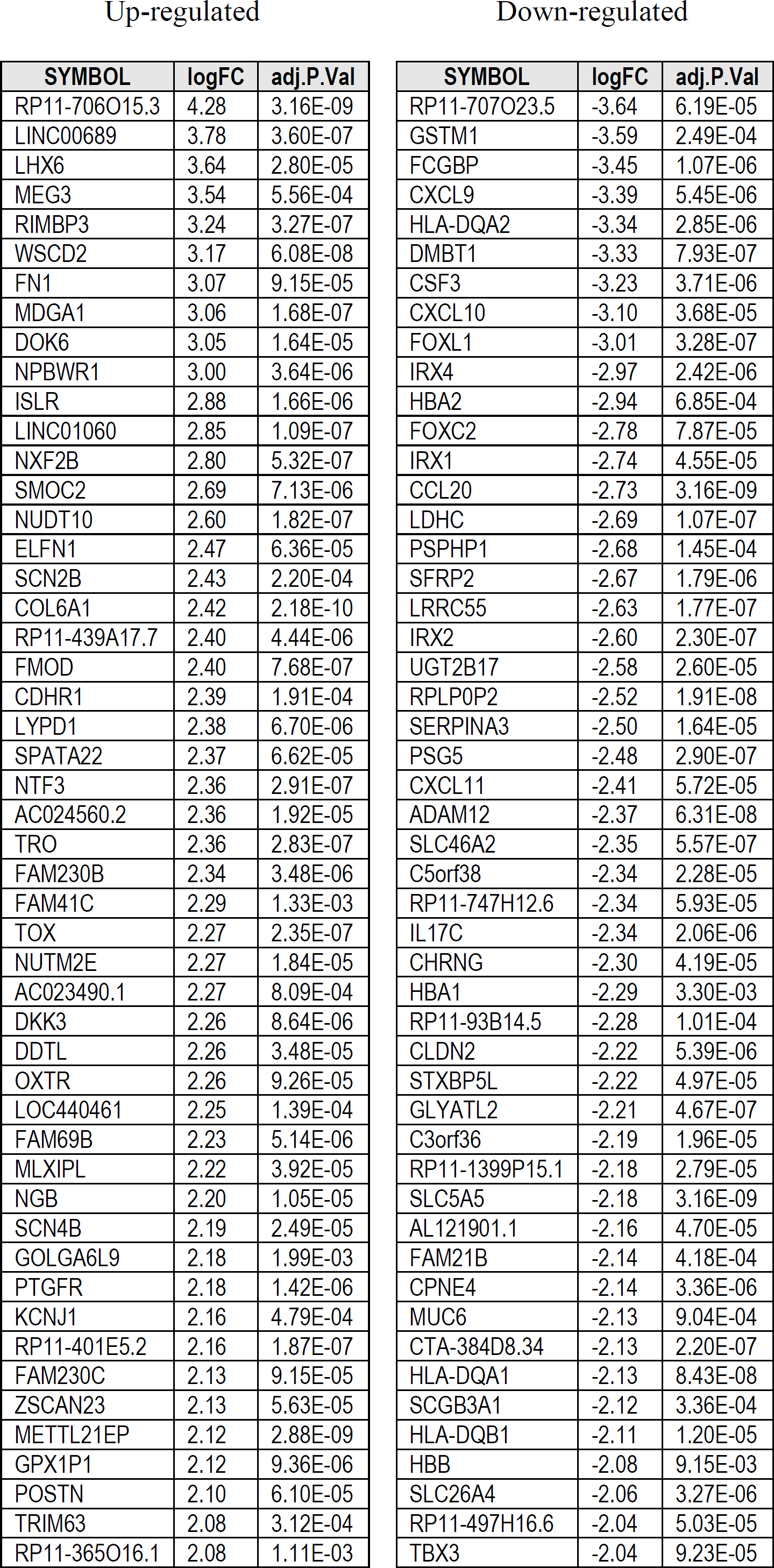
List of differentially expressed genes in baseline vs post-Mepolizumab therapy analysis in all patients. The top 50 most up/down regulated genes are shown.

**Supplementary Table 2.**
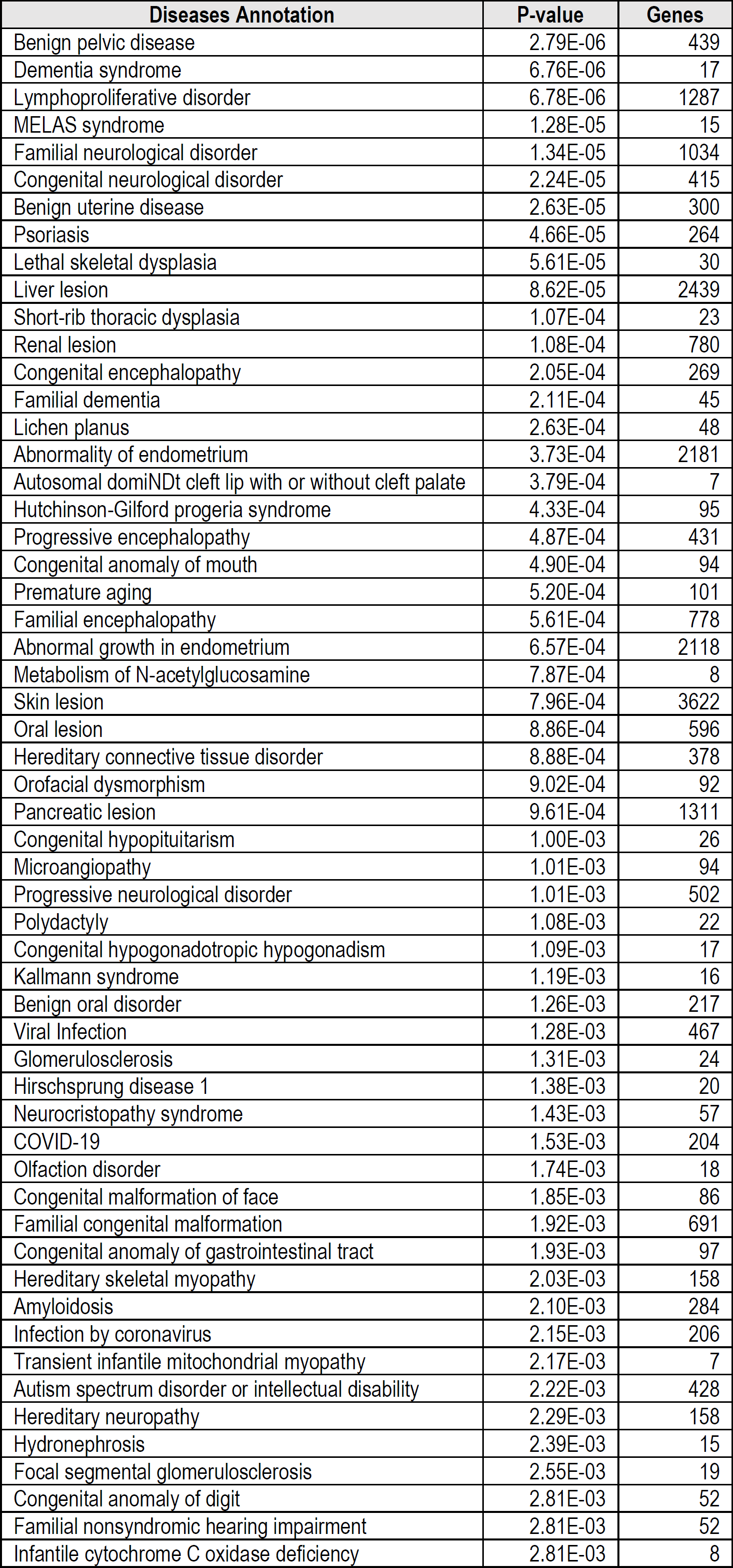
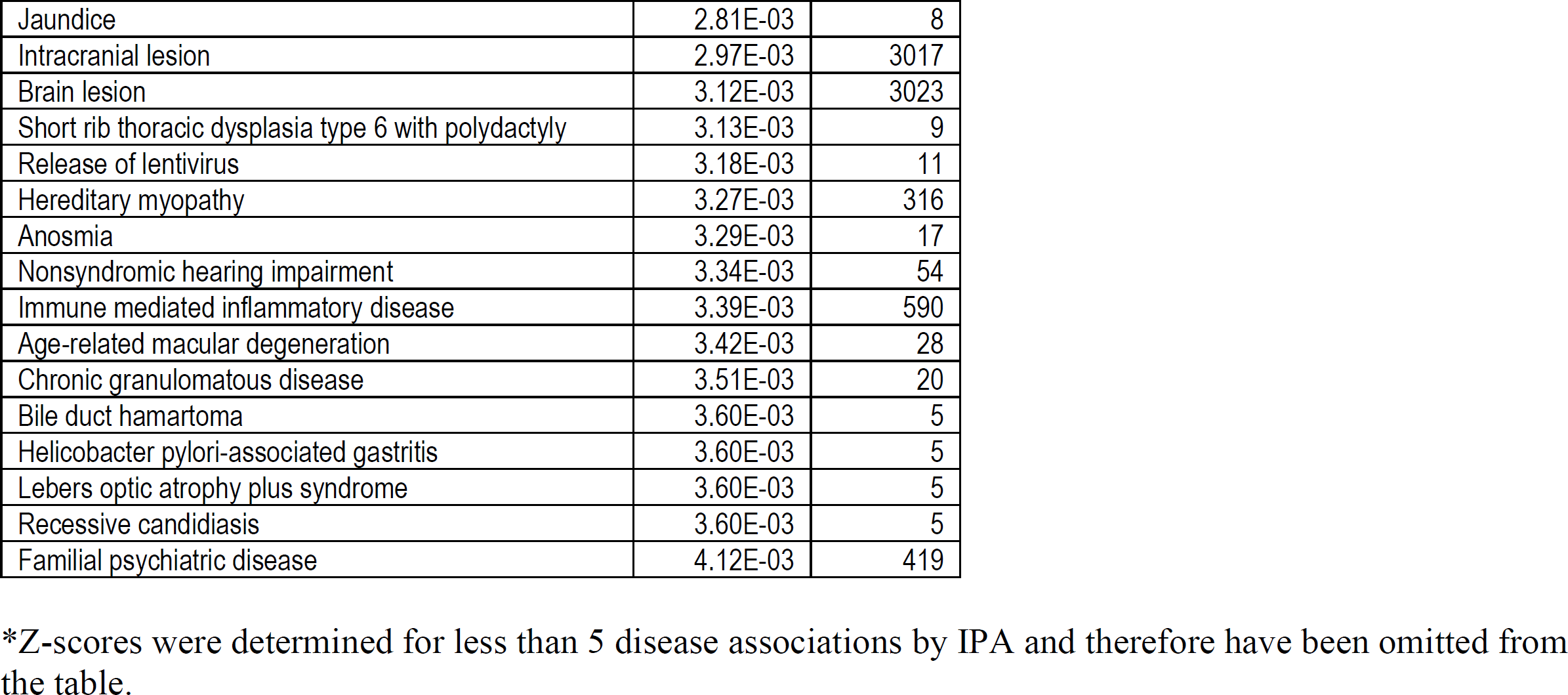
List of disease associations from differential gene expression analysis between baseline vs post-Mepolizumab therapy in all patients*.

**Supplementary Table 3.** Pathway analysis results from differential gene expression analysis between baseline vs post-Mepolizumab therapy in all patients*.

| Ingenuity Canonical Pathways | -log(p-value) | Z-score | Genes |
| --- | --- | --- | --- |
| Antigen Presentation Pathway | 6.33 | ND | 26 |
| iNOS Signaling | 4.41 | -0.85 | 28 |
| Neuroinflammation Signaling Pathway | 3.65 | -0.31 | 114 |
| Xenobiotic Metabolism PXR Signaling Pathway | 3.58 | 2.22 | 73 |
| OX40 Signaling Pathway | 3.56 | 0.91 | 29 |
| Type I Diabetes Mellitus Signaling | 3.25 | -2.27 | 51 |
| Role of IL-17A in Arthritis | 3.24 | ND | 30 |
| Glucocorticoid Receptor Signaling | 3.04 | ND | 184 |
| IL-17A Signaling in Fibroblasts | 2.91 | ND | 21 |
| Ethanol Degradation IV | 2.85 | 1.39 | 13 |
| Putrescine Degradation III | 2.84 | 0.58 | 12 |
| FGF Signaling | 2.76 | 2.67 | 39 |
| Dendritic Cell Maturation | 2.70 | -1.32 | 72 |
| BAG2 Signaling Pathway | 2.57 | 1.00 | 38 |
| IL-17A Signaling in Airway Cells | 2.44 | 0.41 | 32 |
| Lysine Degradation II | 2.44 | 2.24 | 5 |
| Xenobiotic Metabolism Signaling | 2.42 | ND | 103 |
| Sirtuin Signaling Pathway | 2.39 | 1.10 | 109 |
| Notch Signaling | 2.39 | 1.73 | 19 |
| IL-17A Signaling in Gastric Cells | 2.38 | -0.33 | 15 |
| MIF-mediated Glucocorticoid Regulation | 2.33 | -1.00 | 18 |
| Regulation of the Epithelial-Mesenchymal Transition Pathway | 2.28 | ND | 73 |
| Pulmonary Fibrosis Idiopathic Signaling Pathway | 2.27 | 5.09 | 119 |
| Xenobiotic Metabolism AHR Signaling Pathway | 2.26 | 2.26 | 35 |
| Th2 Pathway | 2.25 | 0.71 | 50 |
| ILK Signaling | 2.24 | 2.03 | 75 |
| Hepatic Fibrosis Signaling Pathway | 2.18 | 0.98 | 144 |
| MIF Regulation of Innate Immunity | 2.17 | -0.23 | 21 |
| Histamine Degradation | 2.14 | 1.00 | 9 |
| Crosstalk between Dendritic Cells and Natural Killer Cells | 2.13 | -2.13 | 31 |
| B Cell Development | 2.10 | ND | 14 |
| Role of IL-17F in Allergic Inflammatory Airway Diseases | 2.08 | -0.26 | 22 |
| TNFR2 Signaling | 2.08 | 0.00 | 17 |
| LPS/IL-1 Mediated Inhibition of RXR Function | 2.02 | -0.20 | 80 |
| PCP (Planar Cell Polarity) Pathway | 2.00 | 1.88 | 25 |
| Acetate Conversion to Acetyl-CoA | 1.95 | 0.00 | 4 |
| Epithelial Adherens Junction Signaling | 1.94 | 0.51 | 62 |
| Regulation Of The Epithelial Mesenchymal Transition By Growth Factors Pathway | 1.90 | 2.39 | 72 |
| FAT10 Signaling Pathway | 1.87 | ND | 25 |
| LXR/RXR Activation | 1.85 | -1.51 | 40 |
| Role of Macrophages, Fibroblasts and Endothelial Cells in Rheumatoid Arthritis | 1.84 | ND | 110 |
| Autophagy | 1.82 | -3.58 | 82 |
| Tryptophan Degradation to 2-amino-3-carboxymuconate Semialdehyde | 1.80 | -1.34 | 5 |
| Allograft Rejection Signaling | 1.80 | ND | 20 |
| Xenobiotic Metabolism CAR Signaling Pathway | 1.77 | 2.50 | 64 |
| Regulation Of The Epithelial Mesenchymal Transition In Development Pathway | 1.72 | 2.41 | 33 |
| Tryptophan Degradation X (Mammalian, via Tryptamine) | 1.71 | 0.91 | 11 |
| Ethanol Degradation II | 1.71 | 1.60 | 14 |
| PD-1, PD-L1 cancer immunotherapy pathway | 1.68 | 0.85 | 40 |
| Apelin Pancreas Signaling Pathway | 1.67 | 0.47 | 20 |
| Th1 and Th2 Activation Pathway | 1.67 | ND | 58 |
| Autoimmune Thyroid Disease Signaling | 1.67 | ND | 20 |
| ICOS-ICOSL Signaling in T Helper Cells | 1.64 | -1.18 | 45 |
| Noradrenaline and Adrenaline Degradation | 1.61 | 1.29 | 15 |
| Chemokine Signaling | 1.60 | 0.19 | 31 |
| 4-1BB Signaling in T Lymphocytes | 1.60 | 0.30 | 17 |
| T Cell Receptor Signaling | 1.58 | -1.10 | 69 |
| RANK Signaling in Osteoclasts | 1.57 | 0.54 | 38 |
| PEDF Signaling | 1.56 | 1.51 | 35 |
| Serotonin Degradation | 1.55 | 1.71 | 22 |
| Interferon Signaling | 1.53 | -3.36 | 16 |
| Coronavirus Replication Pathway | 1.53 | 1.89 | 18 |
| Docosahexaenoic Acid (DHA) Signaling | 1.53 | ND | 18 |
| Ferroptosis Signaling Pathway | 1.50 | -2.19 | 48 |
| HER-2 Signaling in Breast Cancer | 1.49 | 2.05 | 83 |
| Serine Biosynthesis | 1.47 | ND | 3 |
| Thiosulfate Disproportionation III (Rhodanese) | 1.47 | ND | 3 |
| FAT10 Cancer Signaling Pathway | 1.44 | 1.41 | 22 |
| CD40 Signaling | 1.42 | 1.23 | 28 |
| Role of BRCA1 in DNA Damage Response | 1.42 | 2.18 | 34 |
| Mitochondrial Dysfunction | 1.42 | ND | 63 |
| Production of Nitric Oxide and Reactive Oxygen Species in Macrophages | 1.41 | -1.13 | 69 |
| Pulmonary Healing Signaling Pathway | 1.41 | 1.81 | 69 |
| Molecular Mechanisms of Cancer | 1.41 | ND | 153 |
| LPS-stimulated MAPK Signaling | 1.40 | -0.18 | 35 |
| WNT/$\beta^2$-catenin Signaling | 1.40 | 0.83 | 60 |
| Dopamine Degradation | 1.40 | 1.16 | 12 |
| NF-κB Signaling | 1.40 | 2.14 | 65 |
| CMP-N-acetylneuraminate Biosynthesis I (Eukaryotes) | 1.39 | -2.00 | 4 |
| Protein Kinase A Signaling | 1.39 | -0.40 | 131 |
| IL-9 Signaling | 1.39 | 0.83 | 16 |
| Lysine Degradation V | 1.39 | 2.00 | 4 |
| Insulin Receptor Signaling | 1.38 | 0.00 | 52 |
| PPAR Signaling | 1.37 | 1.00 | 40 |
| Fatty Acid β-oxidation | 1.37 | 1.00 | 9 |
| Tumor Microenvironment Pathway | 1.36 | 1.92 | 63 |
| Th1 Pathway | 1.35 | 0.00 | 41 |
| Reelin Signaling in Neurons | 1.35 | 1.04 | 54 |
| Endocannabinoid Cancer Inhibition Pathway | 1.34 | -2.77 | 55 |
| Amyloid Processing | 1.34 | 1.16 | 22 |
| Graft-versus-Host Disease Signaling | 1.31 | ND | 20 |
| DNA Double-Strand Break Repair by Homologous Recombination | 1.30 | ND | 8 |

\*Not determined (ND) indicates pathways for which no activity prediction can be made, even though genes are significantly associated with that pathway.

**Supplementary Table 4.** Upstream regulator analysis results from differential gene expression analysis comparing baseline vs post-Mepolizumab therapy in all patients*.

| Upstream Regulator | Molecule Type | Z-score | P-value | Genes |
| --- | --- | --- | --- | --- |
| TNF-alpha | cytokine | -4.381 | 2.13E-09 | 188 |
| LONP1 | peptidase | -2.535 | 1.10E-04 | 14 |
| MT-TE | other | ND | 3.91E-04 | 7 |
| NEUROG1 | transcription regulator | 0.186 | 4.75E-04 | 29 |
| ERK1/2 | group | 0.816 | 5.50E-04 | 39 |
| Interferon alpha | group | -3.257 | 1.05E-03 | 72 |
| mir-183 | microRNA | -0.795 | 1.14E-03 | 9 |
| IL1RAP | transmembrane receptor | -0.816 | 1.20E-03 | 6 |
| IFNG | cytokine | -4.711 | 1.52E-03 | 89 |
| IL13 | cytokine | 1.626 | 1.71E-03 | 79 |
| IL1A | cytokine | -2.597 | 2.15E-03 | 34 |
| PELP1 | other | ND | 2.15E-03 | 34 |
| TGFB1 | growth factor | 1.341 | 2.42E-03 | 85 |
| TLR3 | transmembrane receptor | -1.886 | 2.48E-03 | 33 |
| TP63 | transcription regulator | 1.202 | 2.83E-03 | 95 |
| APP | other | -1.56 | 2.98E-03 | 13 |
| BRD4 | kinase | 1.964 | 3.21E-03 | 24 |
| IL1B | cytokine | -1.75 | 3.33E-03 | 69 |
| VBP1 | other | -2.236 | 3.69E-03 | 5 |
| ERVW-1 | other | -0.246 | 3.69E-03 | 5 |
| SLC9A3R1 | transporter | 0.555 | 3.69E-03 | 5 |
| CG | complex | -1.43 | 3.70E-03 | 77 |
| TGFBR2 | kinase | ND | 3.83E-03 | 30 |
| MYC | transcription regulator | 1.782 | 3.88E-03 | 38 |
| TREM1 | transmembrane receptor | -0.928 | 4.25E-03 | 67 |
| MRTFB | transcription regulator | 2.967 | 5.21E-03 | 30 |
| TREX1 | enzyme | 5.161 | 5.48E-03 | 41 |
| RELA | transcription regulator | -0.276 | 6.49E-03 | 38 |
| IL17F | cytokine | -1.443 | 7.22E-03 | 7 |
| mir-182 | microRNA | 0.478 | 7.22E-03 | 7 |
| TSLP | cytokine | 0.671 | 7.31E-03 | 15 |
| NFKB1 | transcription regulator | -0.037 | 8.06E-03 | 27 |
| IL17A | cytokine | -2.691 | 8.08E-03 | 29 |
| AURK | group | -1.225 | 9.19E-03 | 24 |
| ANLN | other | -1.225 | 9.19E-03 | 24 |
| TEAD4 | transcription regulator | 1.506 | 9.19E-03 | 24 |
| NFkB (complex) | complex | -3.682 | 9.50E-03 | 42 |
| IL1RN | cytokine | 3.499 | 1.05E-02 | 31 |
| CXCL8 | cytokine | -0.098 | 1.07E-02 | 23 |
| LATS2 | kinase | -0.093 | 1.07E-02 | 12 |
| CXCL12 | cytokine | -0.814 | 1.08E-02 | 25 |
| IL36A | cytokine | -2 | 1.13E-02 | 4 |
| MAP2K3 | kinase | -2 | 1.13E-02 | 4 |
| MAP2K6 | kinase | -1.995 | 1.13E-02 | 4 |
| RFXANK | transcription regulator | ND | 1.13E-02 | 4 |
| IL1RL2 | transmembrane receptor | -2 | 1.13E-02 | 4 |
| STAT1 | transcription regulator | -4.537 | 1.14E-02 | 39 |
| LATS1 | kinase | -0.389 | 1.21E-02 | 11 |
| GATA4 | transcription regulator | 1.881 | 1.24E-02 | 22 |
| P38 MAPK | group | -1.657 | 1.27E-02 | 43 |
| FOXC1 | transcription regulator | -4.229 | 1.34E-02 | 41 |
| BTK | kinase | -3.499 | 1.51E-02 | 16 |
| NONO | transcription regulator | -4.609 | 1.54E-02 | 49 |
| mir-23 | microRNA | -1.412 | 1.61E-02 | 5 |
| MALSU1 | other | -2.236 | 1.61E-02 | 5 |
| TEAD3 | transcription regulator | 1.627 | 1.67E-02 | 20 |
| TLR7 | transmembrane receptor | -2.748 | 1.67E-02 | 20 |
| WNT5A | cytokine | -2.326 | 1.72E-02 | 31 |
| miR-218-5p (and other miRNAs w/seed UGUGCUU) | mature microRNA | -0.273 | 1.73E-02 | 7 |
| mir-223 | microRNA | -1.147 | 1.73E-02 | 7 |
| NR3C2 | ligand-dependent nuclear receptor | ND | 1.75E-02 | 6 |
| miR-2392 (miRNAs w/seed AGGAUGG) | mature microRNA | 2.441 | 1.75E-02 | 6 |
| mir-2392 | microRNA | 2.439 | 1.75E-02 | 6 |
| RNASEH2B | other | 5.098 | 1.84E-02 | 37 |
| MUC1 | other | 0.347 | 2.03E-02 | 11 |
| HEIH | other | 1.678 | 2.03E-02 | 14 |
| HNF1A-AS1 | other | 2.137 | 2.03E-02 | 11 |
| PRL | cytokine | -2.107 | 2.06E-02 | 33 |
| CLOCK | transcription regulator | -0.447 | 2.22E-02 | 46 |
| IL33 | cytokine | -0.207 | 2.26E-02 | 18 |
| IRF1 | transcription regulator | -1.342 | 2.33E-02 | 10 |
| GRN | growth factor | -1.462 | 2.55E-02 | 33 |
| OSCAR | other | -1.807 | 2.55E-02 | 15 |
| TEAD2 | transcription regulator | 2.065 | 2.65E-02 | 19 |
| GSK3B | kinase | -1.528 | 2.67E-02 | 9 |
| PGR | ligand-dependent nuclear receptor | 1.269 | 2.67E-02 | 9 |
| PPP2R5C | other | 2.333 | 2.67E-02 | 9 |
| RNA polymerase II | complex | ND | 2.75E-02 | 12 |
| TEAD1 | transcription regulator | 1.506 | 2.96E-02 | 22 |
| TLR9 | transmembrane receptor | -3.446 | 2.96E-02 | 22 |
| MDM4 | transcription regulator | 0.726 | 3.04E-02 | 8 |
| ATF4 | transcription regulator | 1.134 | 3.04E-02 | 8 |
| Nr1h | group | 0.708 | 3.20E-02 | 11 |
| TCR | complex | -0.813 | 3.33E-02 | 79 |
| ILF3 | transcription regulator | 0.232 | 3.33E-02 | 31 |
| Histone h4 | group | ND | 3.43E-02 | 7 |
| miR-23a-3p (and other miRNAs w/seed UCACAUU) | mature microRNA | 0.316 | 3.43E-02 | 7 |
| LINC00963 | other | -1.134 | 3.43E-02 | 7 |
| SPARC | other | -0.503 | 3.43E-02 | 7 |
| PROX1 | transcription regulator | 0.728 | 3.43E-02 | 7 |
| CREBBP | transcription regulator | ND | 3.43E-02 | 7 |
| IKK (complex) | complex | ND | 3.47E-02 | 3 |
| GALNT6 | enzyme | ND | 3.47E-02 | 3 |
| TGM2 | enzyme | ND | 3.47E-02 | 3 |
| CTBP1 | enzyme | ND | 3.47E-02 | 3 |
| EPRS1 | enzyme | ND | 3.47E-02 | 3 |
| SMO | G-protein coupled receptor | ND | 3.47E-02 | 3 |
| Tnf receptor | group | ND | 3.47E-02 | 3 |
| MAP2K7 | kinase | ND | 3.47E-02 | 3 |
| miR-516a-3p (and other miRNAs w/seed GCUUCCU) | mature microRNA | ND | 3.47E-02 | 3 |
| miR-101-3p (and other miRNAs w/seed ACAGUAC) | mature microRNA | ND | 3.47E-02 | 3 |
| ASCL1 | transcription regulator | ND | 3.47E-02 | 3 |
| KDM4A | transcription regulator | ND | 3.47E-02 | 3 |
| CIITA | transcription regulator | ND | 3.47E-02 | 3 |
| RFXAP | transcription regulator | ND | 3.47E-02 | 3 |
| CD74 | transmembrane receptor | ND | 3.47E-02 | 3 |
| SFTPA1 | transporter | ND | 3.47E-02 | 3 |
| FOXO1 | transcription regulator | 1.385 | 3.56E-02 | 32 |
| KRT14 | other | ND | 3.60E-02 | 17 |
| CKAP2L | other | 4.491 | 3.64E-02 | 24 |
| OSM | cytokine | 2.169 | 3.74E-02 | 10 |
| BPIFB1 | other | 0 | 3.74E-02 | 10 |
| EIF4G1 | translation regulator | 0.632 | 3.74E-02 | 10 |
| IL36B | cytokine | -1.78 | 3.81E-02 | 6 |
| PARP1 | enzyme | -0.283 | 3.81E-02 | 6 |
| PGF | growth factor | 2.412 | 3.81E-02 | 6 |
| mir-204 | microRNA | -1.767 | 3.81E-02 | 6 |
| IFNAR2 | transmembrane receptor | -2.449 | 3.81E-02 | 6 |
| SMAD3 | transcription regulator | -0.344 | 3.98E-02 | 25 |
| IRF3 | transcription regulator | -0.65 | 4.08E-02 | 12 |
| Ferritin | complex | 0.555 | 4.12E-02 | 5 |
| IL36G | cytokine | -1.294 | 4.12E-02 | 5 |
| PARK7 | enzyme | 0.13 | 4.12E-02 | 5 |
| FGF18 | growth factor | -0.555 | 4.12E-02 | 5 |
| mir-96 | microRNA | 0.447 | 4.12E-02 | 5 |
| MRPL14 | other | -2.236 | 4.12E-02 | 5 |
| FURIN | peptidase | 1.342 | 4.12E-02 | 5 |
| CUX1 | transcription regulator | -0.447 | 4.12E-02 | 5 |
| TRAF3IP2 | enzyme | -1.964 | 4.18E-02 | 4 |
| SDCBP | enzyme | 0.211 | 4.18E-02 | 4 |
| KDM6B | enzyme | 1 | 4.18E-02 | 4 |
| Ctbp | group | -2 | 4.18E-02 | 4 |
| mir-203 | microRNA | -1.154 | 4.18E-02 | 4 |
| PFDN6 | other | -2 | 4.18E-02 | 4 |
| PFDN2 | other | -2 | 4.18E-02 | 4 |
| LINC01234 | other | -0.927 | 4.18E-02 | 4 |
| IRF7 | transcription regulator | -1.997 | 4.18E-02 | 4 |
| JUNB | transcription regulator | 0.058 | 4.20E-02 | 16 |
| HAND2 | transcription regulator | 1.887 | 4.20E-02 | 14 |
| ERBB2 | kinase | -0.223 | 4.26E-02 | 46 |
| TP53 | transcription regulator | 0.154 | 4.41E-02 | 76 |
| SP1 | transcription regulator | 2.522 | 4.57E-02 | 33 |
| TLR4 | transmembrane receptor | -2.093 | 4.62E-02 | 19 |
| Collagen type II | complex | -1.508 | 4.79E-02 | 11 |
| ADIPOQ | other | -1.577 | 4.79E-02 | 11 |
| EWSR1-FLI1 | fusion gene/product | -2.668 | 4.91E-02 | 31 |

\*Not determined (ND).

**Supplementary Table 5.** Downstream functional effects analysis results from differential gene expression analysis baseline vs post-Mepolizumab therapy in all patients*.

| Functions Annotation | P-value | Z-score | Genes |
| --- | --- | --- | --- |
| Cell movement | 1.96E-06 | -0.75 | 572 |
| Necrosis | 2.96E-06 | 0.60 | 522 |
| Organization of cytoskeleton | 5.53E-06 | 1.84 | 320 |
| Invasion of cells | 6.26E-06 | 0.74 | 383 |
| Microtubule dynamics | 9.37E-06 | 2.04 | 233 |
| Apoptosis | 1.95E-05 | 1.15 | 570 |
| Migration of cells | 2.53E-05 | -0.16 | 508 |
| Necrosis of epithelial tissue | 3.27E-05 | -0.18 | 109 |
| Organization of cytoplasm | 7.31E-05 | 1.84 | 368 |
| Formation of cellular protrusions | 1.69E-04 | 1.90 | 167 |
| Angiogenesis | 2.15E-04 | 0.46 | 260 |
| Transport of monovalent inorganic cation | 2.37E-04 | -0.03 | 61 |
| Cell survival | 2.42E-04 | 1.73 | 284 |
| Transport of metal | 3.00E-04 | -0.52 | 110 |
| Cell viability | 3.36E-04 | 0.96 | 261 |
| Development of vasculature | 4.22E-04 | 0.46 | 262 |
| Cell death of epithelial cells | 4.49E-04 | -0.54 | 69 |
| Transcription | 5.21E-04 | 1.41 | 642 |
| Proliferation of connective tissue cells | 6.61E-04 | -0.86 | 99 |
| Metabolism of N-acetylglucosamine | 7.87E-04 | ND | 8 |
| Monocytopoiesis | 8.22E-04 | -0.63 | 33 |
| Cell proliferation of breast cell lines | 9.19E-04 | 0.13 | 67 |
| Activation of peripheral blood neutrophils | 1.10E-03 | -2.93 | 9 |
| Cell death of breast cell lines | 1.13E-03 | -0.01 | 40 |
| Cell viability of motor neurons | 1.17E-03 | 1.11 | 6 |
| Growth of connective tissue | 1.18E-03 | 0.05 | 122 |
| Transcription of RNA | 1.31E-03 | 1.37 | 564 |
| Unwinding of DNA | 1.36E-03 | ND | 14 |
| Transport of Na<sup>+</sup> | 1.52E-03 | 0.25 | 26 |
| Vasculogenesis | 1.61E-03 | 1.23 | 207 |
| Proliferation of epidermal cells | 1.65E-03 | -0.62 | 33 |
| Transport of inorganic cation | 1.84E-03 | 0.51 | 96 |
| Proliferation of epithelial cells | 1.90E-03 | -0.74 | 77 |
| Apoptosis of epithelial cells | 1.97E-03 | -0.28 | 44 |
| Transport of metal ion | 2.16E-03 | 0.54 | 89 |
| Conformational modification of DNA | 2.16E-03 | ND | 16 |
| Formation of neutrophil extracellular trap | 2.17E-03 | 0.56 | 7 |
| Transport of Cu<sup>2+</sup> | 2.17E-03 | ND | 7 |
| Cell movement of lung cell lines | 2.24E-03 | 2.00 | 20 |
| Transport of cation | 2.31E-03 | 0.51 | 105 |
| Serine phosphorylation | 2.31E-03 | ND | 41 |
| Growth of skin | 2.33E-03 | 0.40 | 39 |
| Myelopoiesis of leukocytes | 2.62E-03 | -0.80 | 42 |
| Serine phosphorylation of protein | 2.81E-03 | ND | 8 |
| Survival of bacteria | 3.04E-03 | 2.07 | 12 |
| Respiratory burst | 3.05E-03 | -0.83 | 21 |
| Proliferation of lung cell lines | 3.05E-03 | 1.50 | 21 |
| Morphology of head | 3.09E-03 | ND | 101 |
| Reorganization of cytoskeleton | 3.11E-03 | 1.23 | 33 |
| Migration of lung cell lines | 3.29E-03 | 1.95 | 17 |
| Expression of RNA | 3.54E-03 | 1.04 | 675 |
| Glucuronidation of lipid | 3.60E-03 | 0.85 | 5 |
| Growth of epithelial tissue | 3.68E-03 | -0.14 | 159 |
| Autophosphorylation of protein | 3.69E-03 | ND | 58 |
| Transcription of DNA | 3.95E-03 | 1.15 | 513 |

\*Not determined (ND)

**Supplementary Table 6.** List of unique differentially expressed genes in baseline vs post-Mepolizumab therapy in separate responder group and corresponding values in non-responder group analysis. 50 most significant genes based on responder analysis.

| Responders |  |  |  |
| --- | --- | --- | --- |
| SYMBOL | logFC | P.Value | adj.P.Val |
| RP11-778D9.12 | 1.27 | 1.59E-07 | 1.31E-04 |
| WDR36 | 0.44 | 4.61E-07 | 2.11E-04 |
| AL354718.1 | 1.59 | 5.29E-07 | 2.32E-04 |
| CLUL1 | 1.47 | 7.69E-07 | 2.82E-04 |
| CTD-2554C21.2 | 1.93 | 9.10E-07 | 2.87E-04 |
| MTND2P28 | -2.32 | 1.06E-06 | 3.06E-04 |
| RP11-497H16.6 | -2.72 | 1.38E-06 | 3.65E-04 |
| ACACA | 0.37 | 1.59E-06 | 3.75E-04 |
| LDOC1L | 0.43 | 1.95E-06 | 4.27E-04 |
| PRKDC | 0.45 | 2.40E-06 | 4.64E-04 |
| AC004870.5 | 1.75 | 3.08E-06 | 5.26E-04 |
| MIB1 | 0.32 | 3.36E-06 | 5.52E-04 |
| C11orf96 | -2.57 | 3.53E-06 | 5.61E-04 |
| MT-RNR2 | -2.31 | 3.76E-06 | 5.93E-04 |
| GPR89B | 1.34 | 4.02E-06 | 6.20E-04 |
| RSPRY1 | 0.28 | 4.29E-06 | 6.31E-04 |
| NDE1 | 0.43 | 4.41E-06 | 6.39E-04 |
| SLC46A1 | 0.66 | 5.07E-06 | 6.89E-04 |
| COL4A3BP | 0.36 | 5.61E-06 | 6.99E-04 |
| RP11-51J9.5 | -0.91 | 5.81E-06 | 7.07E-04 |
| MARVELD2 | 0.42 | 5.95E-06 | 7.12E-04 |
| CAND1 | 0.26 | 6.31E-06 | 7.40E-04 |
| AAK1 | -0.76 | 6.27E-06 | 7.40E-04 |
| FAM208B | 0.34 | 6.60E-06 | 7.61E-04 |
| RP13-467H17.1 | 1.46 | 6.91E-06 | 7.80E-04 |
| GART | 0.26 | 7.33E-06 | 8.04E-04 |
| CD7 | -1.59 | 7.35E-06 | 8.04E-04 |
| KIF3C | 1.04 | 7.51E-06 | 8.14E-04 |
| CAPRIN1 | 0.35 | 8.05E-06 | 8.26E-04 |
| LUZP1 | 0.32 | 8.04E-06 | 8.26E-04 |
| MT-ND1 | -2.15 | 8.03E-06 | 8.26E-04 |
| RP11-391M20.1 | -0.79 | 8.10E-06 | 8.27E-04 |
| PRRC2B | 0.42 | 8.48E-06 | 8.34E-04 |
| PTK2B | 0.34 | 8.67E-06 | 8.37E-04 |
| VAMP2 | -0.40 | 9.17E-06 | 8.48E-04 |
| TOB2 | 0.45 | 1.00E-05 | 8.94E-04 |
| POLR1B | 0.36 | 1.03E-05 | 9.00E-04 |
| MT-CYB | -2.17 | 1.03E-05 | 9.00E-04 |
| MT-CO2 | -2.19 | 1.04E-05 | 9.03E-04 |
| MT-ND2 | -2.13 | 1.05E-05 | 9.04E-04 |
| EEA1 | 0.39 | 1.11E-05 | 9.45E-04 |
| ADNP | 0.27 | 1.22E-05 | 9.85E-04 |
| MT-ND4 | -2.14 | 1.30E-05 | 1.01E-03 |
| ERF | 0.47 | 1.34E-05 | 1.02E-03 |
| PIK3C2A | 0.28 | 1.37E-05 | 1.04E-03 |
| MT-ATP6 | -2.13 | 1.47E-05 | 1.07E-03 |
| KDELC1 | 1.28 | 1.48E-05 | 1.08E-03 |
| GRHL2 | 0.38 | 1.52E-05 | 1.09E-03 |
| MT-CO3 | -2.15 | 1.51E-05 | 1.09E-03 |
| MMP3 | 2.06 | 1.57E-05 | 1.10E-03 |

Down-regulated
| Non-Responders |  |  |  |
| --- | --- | --- | --- |
| SYMBOL | logFC | P.Value | adj.P.Val |
| RP11-778D9.12 | 0.69 | 1.00E-02 | 7.10E-02 |
| WDR36 | 0.27 | 8.16E-03 | 6.33E-02 |
| AL354718.1 | 0.37 | 2.78E-01 | 5.17E-01 |
| CLUL1 | 0.70 | 3.17E-02 | 1.42E-01 |
| CTD-2554C21.2 | 0.31 | 4.41E-01 | 6.70E-01 |
| MTND2P28 | -1.48 | 1.09E-02 | 7.49E-02 |
| RP11-497H16.6 | -1.87 | 5.98E-03 | 5.21E-02 |
| ACACA | 0.09 | 3.28E-01 | 5.69E-01 |
| LDOC1L | 0.26 | 1.30E-02 | 8.34E-02 |
| PRKDC | 0.25 | 2.74E-02 | 1.31E-01 |
| AC004870.5 | 0.68 | 9.35E-02 | 2.73E-01 |
| MIB1 | 0.18 | 3.18E-02 | 1.42E-01 |
| C11orf96 | -1.26 | 5.56E-02 | 1.99E-01 |
| MT-RNR2 | -1.57 | 1.06E-02 | 7.36E-02 |
| GPR89B | 0.45 | 1.50E-01 | 3.61E-01 |
| RSPRY1 | 0.13 | 6.50E-02 | 2.18E-01 |
| NDE1 | 0.29 | 7.63E-03 | 6.08E-02 |
| SLC46A1 | 0.35 | 3.41E-02 | 1.48E-01 |
| COL4A3BP | 0.25 | 8.95E-03 | 6.69E-02 |
| RP11-51J9.5 | -0.50 | 4.40E-02 | 1.73E-01 |
| MARVELD2 | 0.17 | 1.25E-01 | 3.24E-01 |
| CAND1 | 0.16 | 2.12E-02 | 1.12E-01 |
| AAK1 | -0.54 | 9.22E-03 | 6.81E-02 |
| FAM208B | 0.16 | 7.10E-02 | 2.31E-01 |
| RP13-467H17.1 | 0.64 | 7.72E-02 | 2.43E-01 |
| GART | 0.11 | 1.18E-01 | 3.14E-01 |
| CD7 | -1.13 | 9.64E-03 | 6.96E-02 |
| KIF3C | 0.63 | 1.74E-02 | 9.93E-02 |
| CAPRIN1 | 0.26 | 6.46E-03 | 5.46E-02 |
| LUZP1 | 0.18 | 3.26E-02 | 1.45E-01 |
| MT-ND1 | -1.33 | 2.45E-02 | 1.23E-01 |
| RP11-391M20.1 | -0.19 | 3.69E-01 | 6.09E-01 |
| PRRC2B | 0.28 | 1.54E-02 | 9.27E-02 |
| PTK2B | 0.22 | 1.88E-02 | 1.04E-01 |
| VAMP2 | -0.25 | 2.15E-02 | 1.13E-01 |
| TOB2 | 0.24 | 5.01E-02 | 1.87E-01 |
| POLR1B | 0.21 | 2.71E-02 | 1.30E-01 |
| MT-CYB | -1.32 | 2.85E-02 | 1.34E-01 |
| MT-CO2 | -1.36 | 2.58E-02 | 1.26E-01 |
| MT-ND2 | -1.46 | 1.41E-02 | 8.79E-02 |
| EEA1 | 0.22 | 3.40E-02 | 1.48E-01 |
| ADNP | 0.17 | 2.17E-02 | 1.14E-01 |
| MT-ND4 | -1.34 | 2.66E-02 | 1.29E-01 |
| ERF | 0.30 | 1.90E-02 | 1.05E-01 |
| PIK3C2A | 0.16 | 3.54E-02 | 1.52E-01 |
| MT-ATP6 | -1.30 | 3.12E-02 | 1.41E-01 |
| KDELC1 | 0.40 | 2.33E-01 | 4.68E-01 |
| GRHL2 | 0.25 | 1.71E-02 | 9.84E-02 |
| MT-CO3 | -1.31 | 3.19E-02 | 1.43E-01 |
| MMP3 | 1.33 | 1.52E-02 | 9.20E-02 |

Supplementary Table 7. List of unique differentially expressed genes baseline vs post-Mepolizumab therapy in separate non-responder group and corresponding values in responder group analysis. 50 most significant genes based on non-responder analysis.

| Non-Responders |  |  |  |
| --- | --- | --- | --- |
| SYMBOL | logFC | P.Value | adj.P.Val |
| PAPPA | 3.87 | 2.01E-07 | 2.09E-04 |
| CAMK1 | 1.37 | 3.36E-07 | 2.77E-04 |
| BANK1 | -0.74 | 3.39E-06 | 6.88E-04 |
| SOGA2 | 1.45 | 3.45E-06 | 6.88E-04 |
| CDC20B | 1.70 | 4.34E-06 | 7.89E-04 |
| SETD3 | -0.34 | 4.43E-06 | 7.92E-04 |
| GLUL | -0.68 | 4.59E-06 | 7.98E-04 |
| RP11-494M8.4 | 2.70 | 5.09E-06 | 8.24E-04 |
| PCOLCE2 | 3.78 | 6.47E-06 | 9.37E-04 |
| FAM154A | 2.24 | 7.17E-06 | 9.74E-04 |
| LIMS2 | 2.53 | 9.38E-06 | 1.13E-03 |
| NUAK1 | 0.72 | 9.37E-06 | 1.13E-03 |
| AC074011.2 | 3.06 | 9.60E-06 | 1.14E-03 |
| PRKAA2 | -0.74 | 1.11E-05 | 1.26E-03 |
| MOCS2 | -0.37 | 1.12E-05 | 1.26E-03 |
| AC087499.7 | 3.31 | 1.23E-05 | 1.32E-03 |
| RP11-747H12.6 | -3.85 | 1.34E-05 | 1.38E-03 |
| COX14 | -0.65 | 1.46E-05 | 1.44E-03 |
| MPZL1 | -0.51 | 1.55E-05 | 1.50E-03 |
| RP11-553P9.1 | 2.04 | 1.77E-05 | 1.62E-03 |
| HRSP12 | -0.49 | 1.93E-05 | 1.72E-03 |
| RP11-235G24.3 | 3.04 | 2.35E-05 | 1.97E-03 |
| RP11-1149O23.3 | -0.86 | 2.40E-05 | 1.97E-03 |
| RP11-305L7.7 | 1.74 | 2.46E-05 | 2.00E-03 |
| DRAM1 | -0.81 | 2.54E-05 | 2.02E-03 |
| UGT2B17 | -3.85 | 2.68E-05 | 2.09E-03 |
| ARHGEF26 | 0.96 | 2.78E-05 | 2.15E-03 |
| ZNF648 | -1.73 | 3.12E-05 | 2.32E-03 |
| LEPREL1 | 1.25 | 3.34E-05 | 2.39E-03 |
| RP11-44N21.1 | 1.56 | 3.83E-05 | 2.57E-03 |
| ZNHIT6 | -0.71 | 3.85E-05 | 2.58E-03 |
| ZNF833P | 2.05 | 3.86E-05 | 2.58E-03 |
| RP11-707O23.5 | -6.20 | 4.36E-05 | 2.83E-03 |
| CLIC4 | -0.69 | 4.47E-05 | 2.90E-03 |
| CXCL10 | -4.80 | 4.67E-05 | 2.97E-03 |
| USF1 | -0.47 | 4.82E-05 | 3.05E-03 |
| ZNF846 | 0.78 | 5.03E-05 | 3.13E-03 |
| RHOQ | -0.34 | 5.15E-05 | 3.18E-03 |
| RP11-177C12.1 | 1.94 | 5.24E-05 | 3.21E-03 |
| LST1 | -2.15 | 5.36E-05 | 3.25E-03 |
| RP11-488I20.8 | 3.33 | 5.51E-05 | 3.32E-03 |
| TMEM194A | 0.57 | 5.57E-05 | 3.34E-03 |
| RP11-110G21.2 | -1.71 | 5.68E-05 | 3.37E-03 |
| ZDHHC12 | -0.43 | 5.86E-05 | 3.43E-03 |
| AP000704.5 | 1.57 | 5.96E-05 | 3.43E-03 |
| SPATA22 | 3.49 | 6.11E-05 | 3.49E-03 |
| KCNJ15 | -0.69 | 6.80E-05 | 3.75E-03 |
| P4HA3 | 2.69 | 6.99E-05 | 3.81E-03 |
| ETV7 | -1.10 | 7.33E-05 | 3.93E-03 |
| FBXL20 | 0.74 | 7.60E-05 | 4.00E-03 |

Down-regulated
| Responders |  |  |  |
| --- | --- | --- | --- |
| SYMBOL | logFC | P.Value | adj.P.Val |
| PAPPA | 0.65 | 1.26E-01 | 2.88E-01 |
| CAMK1 | 0.42 | 2.24E-02 | 9.84E-02 |
| BANK1 | -0.26 | 2.44E-02 | 1.04E-01 |
| SOGA2 | 0.47 | 3.02E-02 | 1.18E-01 |
| CDC20B | 0.66 | 1.05E-02 | 6.08E-02 |
| SETD3 | -0.08 | 1.20E-01 | 2.80E-01 |
| GLUL | -0.25 | 1.74E-02 | 8.34E-02 |
| RP11-494M8.4 | 0.65 | 1.00E-01 | 2.51E-01 |
| PCOLCE2 | 1.15 | 3.20E-02 | 1.22E-01 |
| FAM154A | 0.57 | 8.51E-02 | 2.27E-01 |
| LIMS2 | 0.67 | 7.66E-02 | 2.13E-01 |
| NUAK1 | 0.24 | 3.92E-02 | 1.38E-01 |
| AC074011.2 | 0.75 | 9.88E-02 | 2.49E-01 |
| PRKAA2 | -0.25 | 4.05E-02 | 1.41E-01 |
| MOCS2 | -0.17 | 8.31E-03 | 5.24E-02 |
| AC087499.7 | 1.15 | 2.87E-02 | 1.14E-01 |
| RP11-747H12.6 | -1.07 | 8.39E-02 | 2.25E-01 |
| COX14 | -0.29 | 9.33E-03 | 5.64E-02 |
| MPZL1 | -0.18 | 4.37E-02 | 1.48E-01 |
| RP11-553P9.1 | 0.55 | 1.04E-01 | 2.57E-01 |
| HRSP12 | -0.14 | 8.80E-02 | 2.32E-01 |
| RP11-235G24.3 | 0.06 | 9.03E-01 | 9.46E-01 |
| RP11-1149O23.3 | -0.33 | 2.93E-02 | 1.16E-01 |
| RP11-305L7.7 | 0.67 | 2.19E-02 | 9.72E-02 |
| DRAM1 | -0.30 | 3.70E-02 | 1.33E-01 |
| UGT2B17 | -1.32 | 4.02E-02 | 1.40E-01 |
| ARHGEF26 | 0.42 | 1.11E-02 | 6.34E-02 |
| ZNF648 | -0.32 | 2.91E-01 | 4.84E-01 |
| LEPREL1 | 0.36 | 9.51E-02 | 2.43E-01 |
| RP11-44N21.1 | 0.64 | 1.97E-02 | 9.07E-02 |
| ZNHIT6 | -0.32 | 1.30E-02 | 6.93E-02 |
| ZNF833P | 0.92 | 1.04E-02 | 6.06E-02 |
| RP11-707O23.5 | -2.34 | 1.57E-02 | 7.84E-02 |
| CLIC4 | -0.12 | 3.22E-01 | 5.15E-01 |
| CXCL10 | -1.97 | 1.36E-02 | 7.14E-02 |
| USF1 | -0.23 | 8.01E-03 | 5.13E-02 |
| ZNF846 | 0.33 | 2.10E-02 | 9.45E-02 |
| RHOQ | -0.09 | 1.44E-01 | 3.13E-01 |
| RP11-177C12.1 | 0.64 | 5.35E-02 | 1.68E-01 |
| LST1 | -0.74 | 5.54E-02 | 1.71E-01 |
| RP11-488I20.8 | 1.04 | 7.40E-02 | 2.08E-01 |
| TMEM194A | 0.28 | 7.79E-03 | 5.05E-02 |
| RP11-110G21.2 | -0.71 | 2.17E-02 | 9.64E-02 |
| ZDHHC12 | -0.20 | 1.22E-02 | 6.67E-02 |
| AP000704.5 | 0.19 | 4.70E-01 | 6.50E-01 |
| SPATA22 | 1.44 | 1.82E-02 | 8.60E-02 |
| KCNJ15 | -0.23 | 7.16E-02 | 2.04E-01 |
| P4HA3 | 0.83 | 4.99E-02 | 1.61E-01 |
| ETV7 | -0.51 | 1.37E-02 | 7.18E-02 |
| FBXL20 | 0.08 | 5.56E-01 | 7.17E-01 |

Supplementary Table 8. List of disease associations from differential gene expression analysis in separate responder or non-responder groups. Due to a large number of disease associations only auto-immune, infectious, inflammatory, or respiratory diseases have been shown.

| Common |  |  |  |  |  |  |
| --- | --- | --- | --- | --- | --- | --- |
| Diseases Annotation | Group | P-value | Genes | Group | P-value | Genes |
| Non-malignant disorder | R | 9.48E-13 | 501 | NR | 6.50E-11 | 373 |
| Benign lesion | R | 1.36E-07 | 259 | NR | 2.21E-06 | 192 |
| Viral Infection | R | 8.41E-07 | 282 | NR | 3.61E-09 | 227 |
| Infection by RNA virus | R | 8.21E-06 | 166 | NR | 1.52E-06 | 131 |
| COVID-19 | R | 9.42E-06 | 129 | NR | 9.18E-07 | 104 |
| Fibrosis | R | 1.39E-04 | 121 | NR | 2.87E-03 | 86 |
| Immune mediated inflammatory disease | R | 1.41E-04 | 334 | NR | 1.48E-05 | 258 |
| Systemic autoimmune syndrome | R | 4.27E-04 | 334 | NR | 1.64E-05 | 261 |
| Inflammation of organ | R | 4.38E-04 | 213 | NR | 2.09E-04 | 163 |
| Eosinophilia | R | 6.55E-04 | 37 | NR | 1.93E-03 | 28 |

| Non-Responders only |  |  |  |
| --- | --- | --- | --- |
| Diseases Annotation | Group | P-value | Genes |
| Psoriasis | NR | 2.84E-06 | 123 |
| Skin lesion | NR | 3.07E-06 | 1407 |
| Diabetes mellitus | NR | 5.90E-06 | 172 |
| Inflammation of muscle | NR | 6.14E-06 | 34 |
| Severe COVID-19 | NR | 1.19E-05 | 40 |
| Lupus erythematosus | NR | 1.36E-05 | 100 |
| Dermatomyositis | NR | 1.44E-05 | 29 |
| Asthma | NR | 1.52E-05 | 52 |
| Systemic lupus erythematosus | NR | 2.68E-05 | 96 |
| Tuberculosis | NR | 4.14E-05 | 42 |
| Acute respiratory disorder | NR | 4.70E-05 | 68 |
| Chronic pulmonary disease | NR | 5.50E-05 | 42 |
| Severe inflammatory disorder | NR | 5.87E-05 | 63 |
| Inflammatory Bowel Disease | NR | 1.45E-04 | 84 |
| Rheumatoid arthritis | NR | 1.76E-04 | 149 |
| Sepsis | NR | 2.32E-04 | 35 |
| Glomerulonephritis | NR | 2.75E-04 | 33 |
| Chronic inflammatory disorder | NR | 2.98E-04 | 217 |
| Viral respiratory infection | NR | 3.08E-04 | 45 |
| Severe pulmonary disease | NR | 3.13E-04 | 47 |
| Chronic obstructive pulmonary disease | NR | 3.31E-04 | 37 |
| Crohn disease | NR | 3.37E-04 | 62 |
| Non-traumatic arthropathy | NR | 4.11E-04 | 175 |
| Severe acute respiratory syndrome | NR | 4.70E-04 | 42 |
| Respiratory distress | NR | 7.40E-04 | 53 |
| Infection of respiratory tract | NR | 7.62E-04 | 51 |
| Rheumatic Disease | NR | 7.68E-04 | 194 |
| Acne | NR | 7.90E-04 | 23 |
| Inflammation of joint | NR | 9.36E-04 | 183 |
| Lower respiratory tract disorder | NR | 9.83E-04 | 60 |
| Insulin-dependent diabetes mellitus | NR | 1.04E-03 | 49 |
| Inflammation of airway | NR | 1.08E-03 | 56 |
| Inflammation of respiratory system component | NR | 1.37E-03 | 65 |
| Lichen planus | NR | 1.50E-03 | 23 |
| Abnormality of skin morphology | NR | 1.67E-03 | 72 |
| Early stage COVID-19 | NR | 2.19E-03 | 3 |
| Immunoglobulin deficiency | NR | 2.40E-03 | 13 |
| Replication of virus | NR | 2.57E-03 | 30 |
| Quantity of virus | NR | 2.94E-03 | 5 |
| Polymyositis | NR | 3.40E-03 | 10 |
| Chronic respiratory disorder | NR | 3.42E-03 | 43 |
| Chronic large plaque psoriasis | NR | 3.62E-03 | 6 |
| Chronic small plaque psoriasis | NR | 3.62E-03 | 6 |
| Dermatitis | NR | 3.77E-03 | 79 |

| Responders only |  |  |  |
| --- | --- | --- | --- |
| Diseases Annotation | Group | P-value | Genes |
| Infection by coronavirus | R | 1.08E-05 | 130 |
| Abdominal lesion | R | 5.03E-05 | 2565 |

Supplementary Table 9. Upstream regulator analysis results from differential gene expression analysis in separate responder (R) or non-responder (NR) groups*.

| Upstream Regulator | Molecule Type | Z-score | P-value | Genes | Group |
| --- | --- | --- | --- | --- | --- |
| TLR3 | transmembrane receptor | -1.30 | 1.75E-06 | 23 | NR |
| IL1B | cytokine | -2.04 | 1.83E-06 | 41 | NR |
| STAT1 | transcription regulator | -4.12 | 1.62E-05 | 26 | NR |
| Interferon alpha | group | -2.12 | 4.26E-05 | 39 | NR |
| TREX1 | enzyme | 4.20 | 9.12E-05 | 25 | NR |
| IL1RN | cytokine | 3.90 | 2.66E-04 | 19 | NR |
| MAP2K3 | kinase | -2.00 | 2.81E-04 | 4 | NR |
| MAP2K6 | kinase | -2.00 | 2.81E-04 | 4 | NR |
| USP8 | peptidase | 3.32 | 4.34E-04 | 11 | NR |
| RNASEH2B | other | 4.26 | 4.71E-04 | 22 | NR |
| TSLP | cytokine | 1.12 | 6.67E-04 | 10 | NR |
| IRF7 | transcription regulator | -2.00 | 1.26E-03 | 4 | NR |
| MYC | transcription regulator | 1.53 | 1.81E-03 | 20 | NR |
| CXCL8 | cytokine | 0.88 | 1.85E-03 | 13 | NR |
| TEAD1 | transcription regulator | 1.96 | 1.89E-03 | 14 | NR |
| TCR | complex | -1.48 | 2.16E-03 | 40 | NR |
| IKK (complex) | complex | ND | 2.17E-03 | 3 | NR |
| KDM4A | transcription regulator | ND | 2.17E-03 | 3 | NR |
| NFIX | transcription regulator | ND | 2.17E-03 | 3 | NR |
| RELB | transcription regulator | ND | 2.17E-03 | 3 | NR |
| SFTPA1 | transporter | ND | 2.17E-03 | 3 | NR |
| PRL | cytokine | -2.51 | 2.29E-03 | 19 | NR |
| PTGS2 | enzyme | -1.81 | 2.38E-03 | 7 | NR |
| STAG2 | other | 3.46 | 2.89E-03 | 12 | NR |
| TEAD2 | transcription regulator | 2.31 | 2.89E-03 | 12 | NR |
| IL4 | cytokine | 2.23 | 3.13E-03 | 21 | NR |
| CHI3L1 | enzyme | ND | 3.33E-03 | 8 | NR |
| CCL2 | cytokine | 0.62 | 3.56E-03 | 11 | NR |
| CAMP | other | -0.50 | 4.36E-03 | 10 | NR |
| CHD7 | enzyme | ND | 5.20E-03 | 5 | NR |
| SNCA | enzyme | ND | 5.79E-03 | 19 | NR |
| FCGR2B | transmembrane receptor | 1.98 | 7.10E-03 | 4 | NR |
| Ferritin | complex | 0.00 | 7.10E-03 | 4 | NR |
| SATB2 | transcription regulator | -0.51 | 7.14E-03 | 10 | NR |
| TLR7/8 | group | -1.49 | 7.14E-03 | 10 | NR |
| RTKN | other | ND | 7.84E-03 | 3 | NR |
| S1PR1 | G-protein coupled receptor | ND | 7.84E-03 | 3 | NR |
| S1PR3 | G-protein coupled receptor | ND | 7.84E-03 | 3 | NR |
| CREBBP | transcription regulator | ND | 8.52E-03 | 5 | NR |
| IKBKE | kinase | -0.45 | 8.52E-03 | 5 | NR |
| NPM1 | transcription regulator | 0.78 | 1.02E-02 | 26 | NR |
| RAG2 | enzyme | 0.54 | 1.05E-02 | 14 | NR |
| RNY3 | other | -2.83 | 1.10E-02 | 8 | NR |
| IL33 | cytokine | 1.27 | 1.11E-02 | 10 | NR |
| SMARCC1 | transcription regulator | ND | 1.27E-02 | 4 | NR |
| VDR | transcription regulator | 0.28 | 1.27E-02 | 4 | NR |
| STAT3 | transcription regulator | -0.90 | 1.30E-02 | 20 | NR |
| SETBP1 | transcription regulator | 0.00 | 1.56E-02 | 16 | NR |
| LIN28B | other | 0.00 | 1.63E-02 | 6 | NR |
| GAPLINC | other | ND | 1.68E-02 | 2 | NR |
| GK | kinase | ND | 1.68E-02 | 2 | NR |
| HISTONE | group | ND | 1.68E-02 | 2 | NR |
| IFI16 | transcription regulator | ND | 1.68E-02 | 2 | NR |
| IRX1 | transcription regulator | ND | 1.68E-02 | 2 | NR |
| MAP2K4 | kinase | ND | 1.68E-02 | 2 | NR |
| MUS81 | enzyme | ND | 1.68E-02 | 2 | NR |
| PML | transcription regulator | ND | 1.68E-02 | 2 | NR |
| RPL11 | other | ND | 1.68E-02 | 2 | NR |
| S1PR2 | G-protein coupled receptor | ND | 1.68E-02 | 2 | NR |
| SRSF2 | transcription regulator | ND | 1.68E-02 | 2 | NR |
| ZBTB7A | transcription regulator | ND | 1.68E-02 | 2 | NR |
| LILRB4 | other | ND | 1.77E-02 | 3 | NR |
| PDE4B | enzyme | ND | 1.77E-02 | 3 | NR |
| TLR9 | transmembrane receptor | -3.29 | 1.86E-02 | 11 | NR |
| CHRNA3 | transmembrane receptor | -0.09 | 2.06E-02 | 4 | NR |
| IFNAR2 | transmembrane receptor | -2.00 | 2.06E-02 | 4 | NR |
| KPNB1 | other | 2.00 | 2.06E-02 | 4 | NR |
| PROX1 | transcription regulator | 0.85 | 2.06E-02 | 4 | NR |
| CD276 | other | 2.45 | 2.18E-02 | 6 | NR |
| KRT14 | other | ND | 2.57E-02 | 9 | NR |
| MAP2K1/2 | group | -1.45 | 2.65E-02 | 5 | NR |
| MMP14 | peptidase | -1.39 | 2.65E-02 | 5 | NR |
| IFNA2 | cytokine | -2.04 | 2.73E-02 | 8 | NR |
| CEBPB | transcription regulator | -0.51 | 2.82E-02 | 10 | NR |
| NRXN1 | transporter | ND | 2.85E-02 | 6 | NR |
| PTPN3 | phosphatase | 1.09 | 2.85E-02 | 6 | NR |
| S100A8 | other | -2.00 | 3.09E-02 | 4 | NR |
| IGF2R | transmembrane receptor | ND | 3.20E-02 | 3 | NR |
| KLF1 | transcription regulator | ND | 3.20E-02 | 3 | NR |
| RASSF8 | other | ND | 3.20E-02 | 3 | NR |
| ZNF100 | transcription regulator | ND | 3.20E-02 | 3 | NR |
| ZNF254 | transcription regulator | ND | 3.20E-02 | 3 | NR |
| ZNF429 | transcription regulator | ND | 3.20E-02 | 3 | NR |
| ZNF43 | transcription regulator | ND | 3.20E-02 | 3 | NR |
| ZNF431 | transcription regulator | ND | 3.20E-02 | 3 | NR |
| ZNF528 | other | ND | 3.20E-02 | 3 | NR |
| ZNF665 | other | ND | 3.20E-02 | 3 | NR |
| ZNF708 | other | ND | 3.20E-02 | 3 | NR |
| ZNF85 | transcription regulator | ND | 3.20E-02 | 3 | NR |
| ZNF91 | transcription regulator | ND | 3.20E-02 | 3 | NR |
| CD36 | transmembrane receptor | -0.85 | 3.64E-02 | 9 | NR |
| TLR7 | transmembrane receptor | -2.33 | 3.64E-02 | 9 | NR |
| FOXC1 | transcription regulator | -3.30 | 3.86E-02 | 18 | NR |
| MRTFA | transcription regulator | 0.45 | 3.98E-02 | 8 | NR |
| GLIS1 | transcription regulator | -0.38 | 4.29E-02 | 7 | NR |
| CD14 | transmembrane receptor | -1.95 | 4.37E-02 | 4 | NR |
| DDIT3 | transcription regulator | -1.10 | 4.37E-02 | 4 | NR |
| PRKCE | kinase | 1.98 | 4.37E-02 | 4 | NR |
| CD44 | other | 0.56 | 4.54E-02 | 6 | NR |
| EZH2 | transcription regulator | 1.97 | 4.58E-02 | 11 | NR |
| CD28 | transmembrane receptor | -1.13 | 4.59E-02 | 14 | NR |
| CCN6 | growth factor | ND | 4.60E-02 | 2 | NR |
| CFLAR | other | ND | 4.60E-02 | 2 | NR |
| CIITA | transcription regulator | ND | 4.60E-02 | 2 | NR |
| Cpla2 | group | ND | 4.60E-02 | 2 | NR |
| CTBP1 | enzyme | ND | 4.60E-02 | 2 | NR |
| GALNT6 | enzyme | ND | 4.60E-02 | 2 | NR |
| LRP1 | transmembrane receptor | ND | 4.60E-02 | 2 | NR |
| MAP2K7 | kinase | ND | 4.60E-02 | 2 | NR |
| MMP12 | peptidase | ND | 4.60E-02 | 2 | NR |
| NLRP3 | other | ND | 4.60E-02 | 2 | NR |
| NOX5 | ion channel | ND | 4.60E-02 | 2 | NR |
| NRP2 | kinase | ND | 4.60E-02 | 2 | NR |
| SUZ12 | enzyme | ND | 4.60E-02 | 2 | NR |
| TGM2 | enzyme | ND | 4.60E-02 | 2 | NR |
| Tnf receptor | group | ND | 4.60E-02 | 2 | NR |
| TNFAIP6 | other | ND | 4.60E-02 | 2 | NR |
| XIAP | enzyme | ND | 4.60E-02 | 2 | NR |
| RNASE2 | enzyme | -1.59 | 4.64E-02 | 5 | NR |
| LONP1 | peptidase | -2.34 | 6.55E-07 | 13 | R |
| MT-TE | other | ND | 5.84E-06 | 7 | R |
| CXCL12 | cytokine | -1.25 | 6.20E-04 | 19 | R |
| mir-2392 | microRNA | 2.44 | 6.57E-04 | 6 | R |
| MALSU1 | other | -2.24 | 9.34E-04 | 5 | R |
| POLRMT | enzyme | -2.24 | 9.34E-04 | 5 | R |
| NFKBIZ | transcription regulator | -1.91 | 9.80E-04 | 7 | R |
| PKM | kinase | 0.44 | 1.99E-03 | 7 | R |
| MRPL14 | other | -2.24 | 2.79E-03 | 5 | R |
| MAP3K7 | kinase | -1.70 | 3.55E-03 | 6 | R |
| PTEN | phosphatase | 0.24 | 5.44E-03 | 8 | R |
| ANLN | other | -1.00 | 5.86E-03 | 16 | R |
| AURK | group | -1.00 | 5.86E-03 | 16 | R |
| PARP1 | enzyme | ND | 6.33E-03 | 5 | R |
| ZEB1 | transcription regulator | 0.29 | 9.81E-03 | 9 | R |
| HSF1 | transcription regulator | 2.45 | 1.00E-02 | 25 | R |
| FGF7 | growth factor | ND | 1.13E-02 | 4 | R |
| ITGAV | transmembrane receptor | -1.96 | 1.13E-02 | 4 | R |
| mir-23 | microRNA | -1.09 | 1.13E-02 | 4 | R |
| GDF2 | growth factor | -0.07 | 1.22E-02 | 5 | R |
| CCL5 | cytokine | -0.83 | 1.50E-02 | 13 | R |
| MRTFA | transcription regulator | 1.24 | 1.50E-02 | 11 | R |
| FOSL1 | transcription regulator | 1.97 | 1.71E-02 | 8 | R |
| ITGA6 | transmembrane receptor | ND | 1.98E-02 | 3 | R |
| OTUD7B | peptidase | ND | 1.98E-02 | 3 | R |
| Raf | group | ND | 1.98E-02 | 3 | R |
| SCAP | other | ND | 1.98E-02 | 3 | R |
| TLR5 | transmembrane receptor | ND | 1.98E-02 | 3 | R |
| TNFSF13B | cytokine | ND | 1.98E-02 | 3 | R |
| BCL3 | transcription regulator | -1.17 | 2.08E-02 | 5 | R |
| FURIN | peptidase | 0.00 | 2.26E-02 | 4 | R |
| ITGB3 | transmembrane receptor | -1.00 | 2.26E-02 | 4 | R |
| TP63 | transcription regulator | -0.19 | 2.93E-02 | 51 | R |
| AKT2 | kinase | ND | 3.20E-02 | 2 | R |
| CTBP2 | transcription regulator | ND | 3.20E-02 | 2 | R |
| GAS6 | growth factor | ND | 3.20E-02 | 2 | R |
| GATAD2B | transcription regulator | ND | 3.20E-02 | 2 | R |
| IL1R2 | transmembrane receptor | ND | 3.20E-02 | 2 | R |
| IQGAP1 | other | ND | 3.20E-02 | 2 | R |
| LILRB1 | transmembrane receptor | ND | 3.20E-02 | 2 | R |
| MAPKAPK2 | kinase | ND | 3.20E-02 | 2 | R |
| mir-26 | microRNA | ND | 3.20E-02 | 2 | R |
| NCOR1 | transcription regulator | ND | 3.20E-02 | 2 | R |
| NEDD4 | enzyme | ND | 3.20E-02 | 2 | R |
| NFE2L1 | transcription regulator | ND | 3.20E-02 | 2 | R |
| PTGER4 | G-protein coupled receptor | ND | 3.20E-02 | 2 | R |
| RFX5 | transcription regulator | ND | 3.20E-02 | 2 | R |
| SCGB1A1 | cytokine | ND | 3.20E-02 | 2 | R |
| TP53 | transcription regulator | 0.50 | 3.54E-02 | 44 | R |
| LGALS3 | other | 1.07 | 3.89E-02 | 4 | R |
| NAMPT | cytokine | -0.83 | 3.89E-02 | 4 | R |
| NR3C2 | ligand-dependent nuclear receptor | ND | 3.89E-02 | 4 | R |
| SMAD1 | transcription regulator | -1.98 | 3.89E-02 | 4 | R |
| DUX4 | transcription regulator | -0.43 | 3.98E-02 | 9 | R |
| ARID1A | transcription regulator | 0.82 | 3.99E-02 | 32 | R |
| HAND1 | transcription regulator | ND | 4.30E-02 | 3 | R |
| IL22RA1 | transmembrane receptor | ND | 4.30E-02 | 3 | R |
| KIF26A | other | ND | 4.30E-02 | 3 | R |
| mir-148 | microRNA | ND | 4.30E-02 | 3 | R |
| PIK3R1 | kinase | ND | 4.30E-02 | 3 | R |
| STK4 | kinase | ND | 4.30E-02 | 3 | R |
| TFAP2A | transcription regulator | ND | 4.30E-02 | 3 | R |
| TLR6 | transmembrane receptor | ND | 4.30E-02 | 3 | R |
| VIM | other | ND | 4.30E-02 | 3 | R |
| TGFBR2 | kinase | 0.00 | 4.66E-02 | 19 | R |
| FBN1 | other | -1.25 | 4.78E-02 | 5 | R |
| AGO2 | translation regulator | 2.44 | 5.13E-02 | 6 | R |

\*Not determined (ND)

Supplementary Table 10. Downstream functional effects analysis results from differential gene expression analysis in separate responder (R) or non-responder (NR) groups*.

| Functional Annotation | P-value | Z-score | Genes | Group |
| --- | --- | --- | --- | --- |
| Binding of blood cells | 1.44E-05 | -0.55 | 54 | NR |
| Movement of CD4+ T-lymphocytes | 2.43E-05 | 0.00 | 8 | NR |
| Adhesion of blood cells | 3.24E-05 | -0.70 | 48 | NR |
| Transport of monosaccharide | 3.98E-05 | -0.08 | 20 | NR |
| Chemotaxis | 8.54E-05 | -0.68 | 67 | NR |
| Cell viability of granulocytes | 9.40E-05 | 0.85 | 9 | NR |
| Transport of carbohydrate | 9.54E-05 | -0.54 | 25 | NR |
| Transport of hexose | 1.25E-04 | -0.36 | 18 | NR |
| Apoptosis of B lymphoblastoid cell lines | 1.26E-04 | -2.01 | 7 | NR |
| Cell spreading | 1.32E-04 | 0.35 | 27 | NR |
| Cell death of immune cells | 1.86E-04 | 0.50 | 53 | NR |
| Migration of CD4+ T-lymphocytes | 1.97E-04 | -1.34 | 5 | NR |
| Cell movement of endothelial cell lines | 1.98E-04 | -0.77 | 26 | NR |
| Adhesion of immune cells | 2.26E-04 | -0.30 | 41 | NR |
| Phosphorylation of protein | 2.27E-04 | -1.40 | 98 | NR |
| Serine phosphorylation | 2.32E-04 | 0.56 | 35 | NR |
| Binding of leukocytes | 2.71E-04 | -0.88 | 44 | NR |
| Cell death of blood cells | 2.78E-04 | 0.48 | 55 | NR |
| Activation of cells | 2.89E-04 | -0.57 | 76 | NR |
| Cell viability | 3.23E-04 | -1.09 | 118 | NR |
| Cell survival | 3.41E-04 | -0.35 | 127 | NR |
| Apoptosis of lymphatic system cells | 3.48E-04 | 0.55 | 28 | NR |
| Transport of D-glucose | 3.60E-04 | -0.36 | 15 | NR |
| Inflammatory response | 3.67E-04 | -0.35 | 73 | NR |
| Survival of neural cells | 3.92E-04 | 1.22 | 16 | NR |
| Cell viability of myeloid cells | 4.08E-04 | 1.64 | 12 | NR |
| Cell movement of epithelial cells | 4.17E-04 | -0.28 | 21 | NR |
| Cell viability of leukocytes | 4.25E-04 | 0.53 | 20 | NR |
| Cellular homeostasis | 4.60E-04 | -2.05 | 145 | NR |
| Cytotoxicity of cytotoxic T cells | 5.18E-04 | -2.61 | 7 | NR |
| Morphology of tumor cell lines | 5.29E-04 | ND | 19 | NR |
| Protein kinase cascade | 5.56E-04 | -0.73 | 63 | NR |
| Phosphorylation of L-amino acid | 5.91E-04 | 0.97 | 29 | NR |
| Apoptosis of lymphocytes | 5.98E-04 | 0.13 | 27 | NR |
| Transport of monovalent inorganic cation | 5.98E-04 | 0.60 | 27 | NR |
| Shape change of vascular endothelial cells | 6.09E-04 | -0.31 | 15 | NR |
| Cell movement of endothelial cells | 6.25E-04 | 0.08 | 65 | NR |
| Migration of skin cell lines | 6.32E-04 | -0.39 | 11 | NR |
| Binding of myeloid cells | 7.52E-04 | -0.29 | 26 | NR |
| Cell death of lymphocytes | 7.54E-04 | 0.34 | 31 | NR |
| Interaction of carbohydrate | 8.78E-04 | -1.74 | 13 | NR |
| Proliferation of neuronal cells | 9.64E-04 | 1.96 | 19 | NR |
| Proliferation of lymphoblastoid cell lines | 1.01E-03 | 0.45 | 16 | NR |
| Cell spreading of endothelial cells | 1.05E-03 | -0.43 | 9 | NR |
| Phosphorylation of peptide | 1.08E-03 | 0.98 | 35 | NR |
| Apoptosis of tumor cell lines | 1.14E-03 | -1.39 | 81 | NR |
| Sprouting of vascular endothelial cells | 1.21E-03 | 0.55 | 11 | NR |
| Apoptosis of leukocytes | 1.24E-03 | -0.05 | 38 | NR |
| Catabolism of L-tryptophan | 1.27E-03 | ND | 4 | NR |
| Clustering of cells | 1.27E-03 | ND | 4 | NR |
| Formation of hemidesmosomes | 1.27E-03 | 0.00 | 4 | NR |
| Migration of endothelial cell lines | 1.33E-03 | -0.63 | 20 | NR |
| Import of D-glucose | 1.38E-03 | -0.63 | 10 | NR |
| Metabolism of peptide | 1.47E-03 | 1.15 | 44 | NR |
| Cell death of T lymphocytes | 1.50E-03 | 0.50 | 23 | NR |
| Transport of inorganic cation | 1.51E-03 | 1.28 | 42 | NR |
| Apoptosis of T lymphocytes | 1.58E-03 | 0.10 | 20 | NR |
| Transport of metal ion | 1.70E-03 | 1.71 | 39 | NR |
| Interaction of protein | 1.75E-03 | 1.07 | 21 | NR |
| Binding of granulocytes | 1.76E-03 | -0.09 | 18 | NR |
| Binding of carbohydrate | 1.80E-03 | -1.31 | 12 | NR |
| Serine phosphorylation of peptide | 1.83E-03 | 0.80 | 31 | NR |
| Cell-cell contact | 1.86E-03 | 0.48 | 51 | NR |
| Response of phagocytes | 1.90E-03 | -0.86 | 16 | NR |
| Cell movement of dermal cells | 1.94E-03 | -0.73 | 15 | NR |
| Growth of neurites | 1.94E-03 | 2.06 | 15 | NR |
| Migration of epithelial cells | 1.94E-03 | 0.13 | 15 | NR |
| Outgrowth of neurites | 1.94E-03 | 1.87 | 14 | NR |
| Stimulation of cells | 2.11E-03 | -0.72 | 18 | NR |
| Proliferation of neural cells | 2.15E-03 | 2.06 | 24 | NR |
| Internalization of carbohydrate | 2.16E-03 | -0.11 | 11 | NR |
| Density of microvessel | 2.19E-03 | ND | 3 | NR |
| Migration of cerebral cortex cells | 2.19E-03 | ND | 3 | NR |
| Migration of helper T lymphocytes | 2.19E-03 | ND | 3 | NR |
| Mutagenesis of cells | 2.19E-03 | ND | 3 | NR |
| Parturition | 2.19E-03 | ND | 3 | NR |
| Apoptosis of blood cells | 2.30E-03 | -0.23 | 39 | NR |
| Branching of cells | 2.31E-03 | 1.20 | 16 | NR |
| I-kappaB kinase/NF-kappaB cascade | 2.34E-03 | -1.64 | 26 | NR |
| Migration of thyroid tumor cell lines | 2.36E-03 | 0.82 | 8 | NR |
| Cell movement of skin cell lines | 2.40E-03 | -1.29 | 13 | NR |
| Cell movement of keratinocytes | 2.41E-03 | -1.12 | 14 | NR |
| Formation of gland | 2.42E-03 | ND | 7 | NR |
| Transmigration of cells | 2.59E-03 | -0.86 | 20 | NR |
| Cell viability of squamous cell carcinoma cell lines | 2.82E-03 | 1.29 | 11 | NR |
| Synthesis of DNA | 2.86E-03 | 1.13 | 37 | NR |
| Modification of peptide | 2.88E-03 | 0.98 | 36 | NR |
| Activation of CD4+ T-lymphocytes | 2.94E-03 | ND | 5 | NR |
| Killing of lymphocytes | 2.94E-03 | 0.45 | 5 | NR |
| Morphogenesis of embryonic tissue | 2.94E-03 | ND | 5 | NR |
| Binding of polysaccharide | 2.99E-03 | -1.07 | 9 | NR |
| Transport of metal | 3.00E-03 | 0.96 | 45 | NR |
| Outgrowth of cells | 3.36E-03 | 1.78 | 16 | NR |
| Development of fibroblasts | 3.38E-03 | 1.52 | 8 | NR |
| Transmembrane receptor protein tyrosine kinase signaling pathway | 3.38E-03 | ND | 8 | NR |
| Apoptosis of lymphoblastoid cell lines | 3.40E-03 | -2.40 | 10 | NR |
| Migration of neural cells | 3.40E-03 | -0.89 | 10 | NR |
| Apoptosis of schwannoma cell lines | 3.42E-03 | -1.12 | 4 | NR |
| Interphase of skin cancer cell lines | 3.42E-03 | ND | 4 | NR |
| Migration of central nervous system cells | 3.42E-03 | ND | 4 | NR |
| Immune response of phagocytes | 3.53E-03 | -0.63 | 15 | NR |
| Response of myeloid cells | 3.53E-03 | -0.56 | 15 | NR |
| Hematopoiesis of mononuclear leukocytes | 3.55E-03 | -1.60 | 35 | NR |
| Intercellular communication | 3.57E-03 |  | 23 | NR |
| Colony formation of epithelial cell lines | 3.62E-03 | -0.39 | 6 | NR |
| Formation of mammary gland | 3.62E-03 | ND | 6 | NR |
| Synthesis of nucleoside diphosphates | 3.62E-03 | ND | 6 | NR |
| Binding of blood platelets | 3.64E-03 | 0.97 | 11 | NR |
| Branching of epithelial tissue | 3.74E-03 | 0.90 | 13 | NR |
| Invasion of tumor cell lines | 1.02E-07 | 2.16 | 206 | R |
| Cell movement of tumor cell lines | 1.44E-06 | 1.85 | 124 | R |
| Migration of tumor cell lines | 4.58E-06 | 2.13 | 114 | R |
| Invasion of carcinoma cell lines | 5.07E-06 | 1.73 | 82 | R |
| Activation of peripheral blood neutrophils | 7.27E-06 | -2.93 | 9 | R |
| Organization of cytoskeleton | 8.21E-06 | 1.37 | 189 | R |
| Transcription | 2.02E-05 | 2.34 | 370 | R |
| Invasion of breast cancer cell lines | 4.99E-05 | 1.60 | 65 | R |
| Expression of RNA | 7.88E-05 | 1.96 | 390 | R |
| Microtubule dynamics | 8.20E-05 | 1.46 | 136 | R |
| Organization of cytoplasm | 1.12E-04 | 1.37 | 215 | R |
| Interaction of epithelial cells | 1.67E-04 | 1.39 | 21 | R |
| Hydrolysis of nucleotide | 2.13E-04 | 1.82 | 18 | R |
| Cell movement of fibrosarcoma cell lines | 2.79E-04 | 2.02 | 20 | R |
| Cell death of epithelial cells | 3.03E-04 | -1.16 | 46 | R |
| Invasion of thyroid tumor cell lines | 3.59E-04 | 0.33 | 10 | R |
| Synthesis of nucleotide | 4.15E-04 | -0.59 | 43 | R |
| Migration of fibrosarcoma cell lines | 4.18E-04 | 1.89 | 18 | R |
| Reorganization of cytoskeleton | 4.52E-04 | 1.24 | 23 | R |
| Formation of nuclear foci | 5.83E-04 | -2.53 | 23 | R |
| Apoptosis of epithelial cells | 5.89E-04 | -1.22 | 29 | R |
| Detachment of cells | 6.36E-04 | -0.46 | 10 | R |
| Formation of cellular protrusions | 7.05E-04 | 1.53 | 98 | R |
| Quantity of superoxide | 9.38E-04 | -2.24 | 5 | R |
| Transport of heavy metal | 9.48E-04 | -1.13 | 24 | R |
| Degradation of hydrogen peroxide | 9.85E-04 | ND | 7 | R |

\*Not determined (ND)

Supplementary Table 11. List of differentially methylated CpG sites baseline vs post-Mepolizumab therapy in all patients.

| CpG site | logFC | adj.P.Val | Location (Genome 37) | Annotated gene |
| --- | --- | --- | --- | --- |
| cg09261794 | 0.0274 | 1.06E-02 | Chromosome 6 - 33147897 | COL11A2 |
| cg16243351 | 0.0134 | 1.59E-02 | Chromosome 11 - 35453590 | PAMR1 |
| cg17748184 | 0.0045 | 1.64E-02 | Chromosome 1 - 222639024 | - |
| cg15733917 | -0.0078 | 1.68E-02 | Chromosome 2 - 109150694 | - |
| cg13551227 | 0.0120 | 1.69E-02 | Chromosome 10 - 103604019 | KCNIP2 |
| cg07252627 | 0.0145 | 2.10E-02 | Chromosome 19 - 18378899 | KIAA1683 |
| cg11298262 | 0.0107 | 2.10E-02 | Chromosome 20 - 56009992 | - |
| cg22681114 | 0.0271 | 2.10E-02 | Chromosome 6 - 31123764 | CCHCR1 |
| cg06788631 | 0.0079 | 2.26E-02 | Chromosome 10 - 101300607 | - |
| cg05214404 | 0.0060 | 2.32E-02 | Chromosome 4 - 37455362 | C4orf19 |
| cg09715562 | 0.0411 | 2.32E-02 | Chromosome 22 - 19072685 | DGCR2 |
| cg10648252 | 0.0125 | 2.32E-02 | Chromosome 7 - 101469088 | CUX1 |
| cg18471664 | 0.0065 | 2.32E-02 | Chromosome 3 - 5028143 | - |
| cg14080745 | 0.0045 | 2.44E-02 | Chromosome 20 - 61556425 | DIDO1 |
| cg24669932 | 0.0109 | 2.66E-02 | Chromosome 1 - 8877431 | RERE |
| cg20411278 | 0.0200 | 3.29E-02 | Chromosome 17 - 45728574 | KPNB1 |
| cg24670497 | 0.0113 | 3.29E-02 | Chromosome 1 - 16068127 | TMEM82 |
| cg18301742 | -0.0038 | 3.31E-02 | Chromosome 21 - 34100210 | PAXBP1-AS1 |
| cg05683632 | 0.0130 | 3.67E-02 | Chromosome 19 - 41253983 | C19orf54 |
| cg09423080 | 0.0018 | 3.67E-02 | Chromosome 13 - 76111751 | COMMD6 |
| cg26066560 | 0.0683 | 3.67E-02 | Chromosome 7 - 2086333 | MAD1L1 |
| cg02151918 | 0.0165 | 3.71E-02 | Chromosome 11 - 70211268 | PPFIA1 |
| cg02635677 | 0.0368 | 3.71E-02 | Chromosome 5 - 176823320 | SLC34A1 |
| cg03010429 | 0.0163 | 3.71E-02 | Chromosome 2 - 134352494 | - |
| cg05002732 | 0.0089 | 3.71E-02 | Chromosome 11 - 19262474 | E2F8 |
| cg05095252 | 0.0444 | 3.71E-02 | Chromosome 2 - 150308042 | LYPD6 |
| cg06211272 | 0.0033 | 3.71E-02 | Chromosome 1 - 59012675 | OMA1 |
| cg06301443 | 0.0094 | 3.71E-02 | Chromosome 22 - 19807266 | GNB1L |
| cg08369629 | 0.1184 | 3.71E-02 | Chromosome 8 - 53598501 | RB1CC1 |
| cg09099761 | 0.0192 | 3.71E-02 | Chromosome 10 - 72081690 | LRRC20 |
| cg13469617 | 0.0243 | 3.71E-02 | Chromosome 21 - 40119488 | LINC00114 |
| cg16226507 | 0.0082 | 3.71E-02 | Chromosome 5 - 180432814 | BTNL3 |
| cg16373535 | 0.0176 | 3.71E-02 | Chromosome 5 - 1923122 | - |
| cg17422500 | 0.0031 | 3.71E-02 | Chromosome 1 - 249152952 | ZNF692 |
| cg18663341 | 0.0121 | 3.71E-02 | Chromosome 7 - 45148416 | TBRG4 |
| cg18900832 | 0.0335 | 3.71E-02 | Chromosome 15 - 25303214 | SNORD116-4 |
| cg18918010 | 0.0105 | 3.71E-02 | Chromosome 8 - 123871905 | ZHX2 |
| cg20256649 | 0.0173 | 3.71E-02 | Chromosome 2 - 1941523 | MYT1L |
| cg21136271 | 0.0132 | 3.71E-02 | Chromosome 15 - 93146384 | - |
| cg22347697 | 0.0319 | 3.71E-02 | Chromosome 19 - 47827163 | - |
| cg23406506 | 0.0047 | 3.71E-02 | Chromosome 4 - 110736542 | GAR1 |
| cg27406678 | 0.0104 | 3.71E-02 | Chromosome 4 - 85417612 | NKX6-1 |
| cg24714082 | 0.0241 | 3.90E-02 | Chromosome 1 - 167595191 | - |
| ch.7.3189261R | 0.0080 | 3.90E-02 | Chromosome 7 - 151489100 | PRKAG2 |
| cg18427648 | 0.0050 | 3.92E-02 | Chromosome 4 - 146403869 | SMAD1 |
| cg16827359 | 0.0219 | 4.21E-02 | Chromosome 22 - 49554562 | - |
| cg05579339 | 0.0040 | 4.54E-02 | Chromosome 14 - 78226979 | C14orf178, SNW1 |
| cg07906473 | 0.0154 | 4.54E-02 | Chromosome 16 - 88573286 | ZFPM1 |
| cg12823530 | 0.0158 | 4.54E-02 | Chromosome 14 - 24551984 | NRL |
| cg14876117 | 0.0739 | 4.54E-02 | Chromosome 14 - 51428973 | - |
| cg20207310 | 0.0070 | 4.54E-02 | Chromosome 1 - 244614789 | ADSS |
| cg26744946 | 0.0226 | 4.54E-02 | Chromosome 8 - 145065833 | GRINA |
| cg27333547 | 0.0218 | 4.54E-02 | Chromosome 16 - 88106229 | BANP |

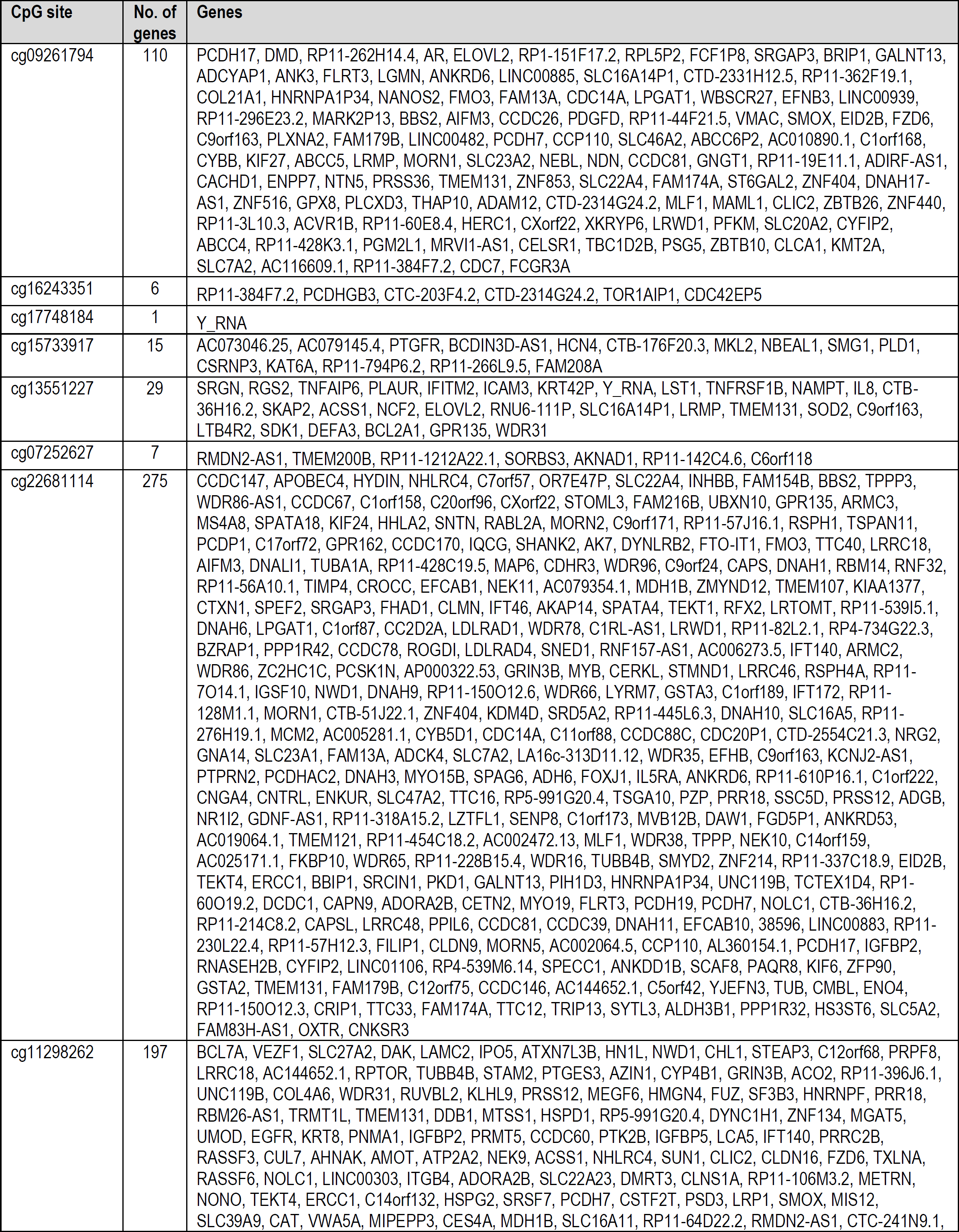
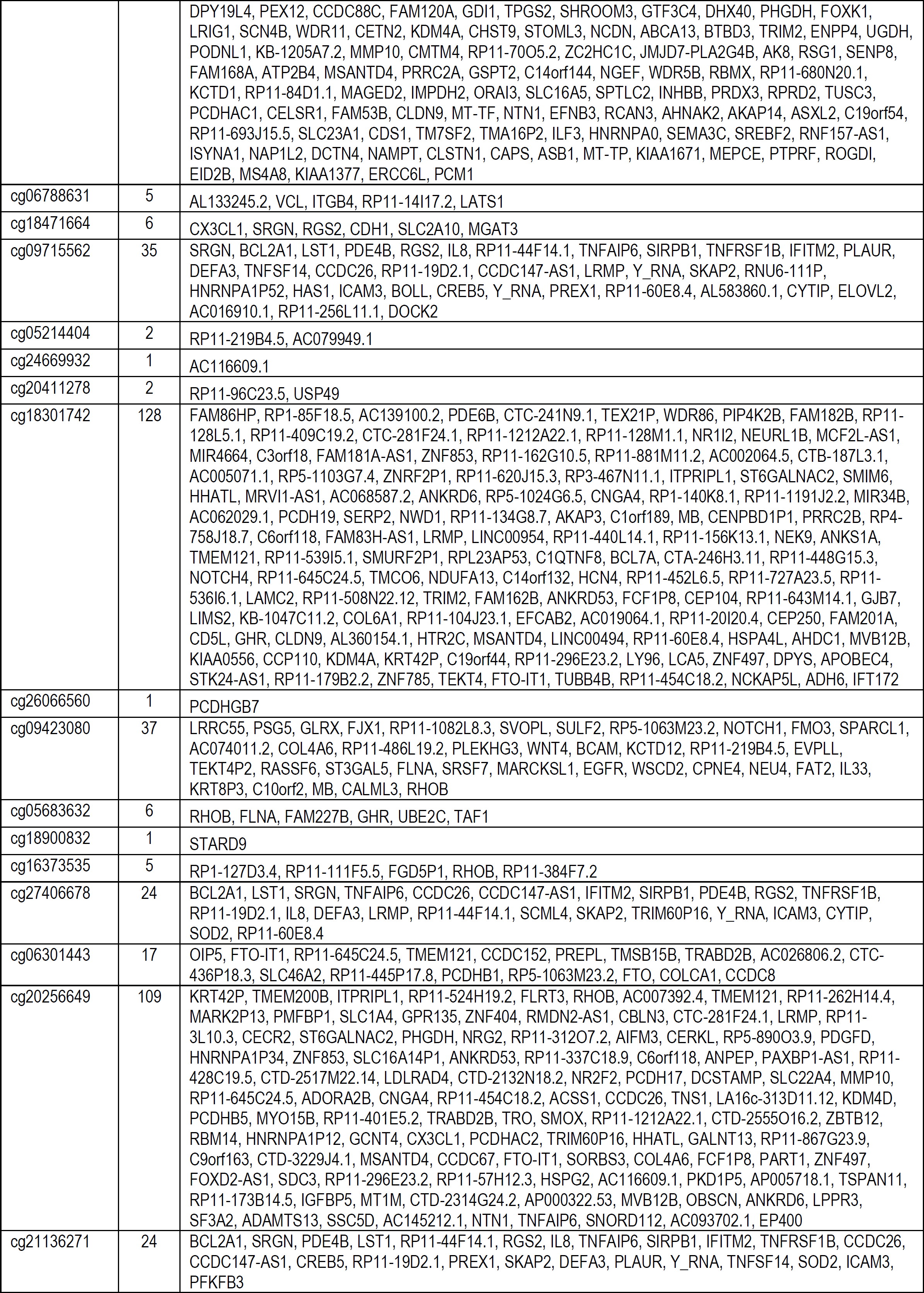
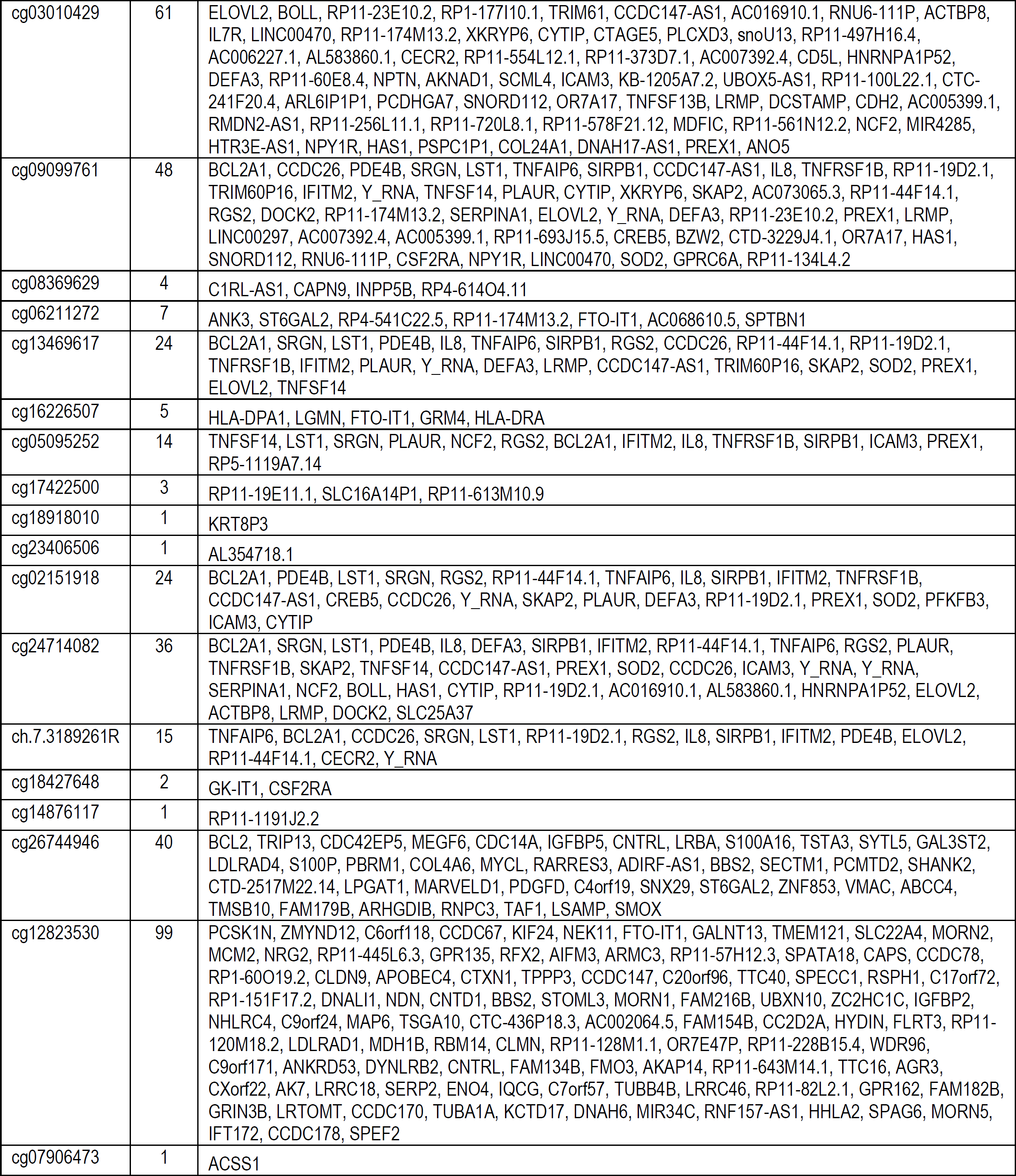
Supplementary Table 12. List of correlated differentially methylated CpG sites and differentially expressed genes from the baseline vs post-Mepolizumab therapy in all patients. List show the CpG site and genes it correlates with.

Supplementary Table 13. List of disease associations from genes identified as part of the differential gene expression and methylation correlation analysis. There were over 500 associations and therefore only those related to the respiratory system/lung are shown for clarity.

| Diseases Annotation | P-value | Genes |
| --- | --- | --- |
| Primary ciliary dyskinesia | 4.33E-09 | 13 |
| Nonsquamous non-small cell lung carcinoma | 3.66E-06 | 125 |
| Small cell lung carcinoma | 5.20E-06 | 138 |
| Lung adenocarcinoma | 7.78E-06 | 123 |
| Malignant neoplasm of respiratory system | 1.26E-05 | 257 |
| Lung carcinoma | 1.42E-05 | 230 |
| Lung cancer | 1.58E-05 | 235 |
| Non-small cell lung carcinoma | 8.64E-05 | 152 |
| Large cell lung carcinoma | 5.78E-04 | 12 |
| Laryngeal squamous cell carcinoma | 6.18E-04 | 39 |
| Non-small cell lung adenosquamous carcinoma | 1.10E-03 | 6 |
| Chronic familial respiratory disorder | 1.14E-03 | 5 |
| Recurrent large cell lung carcinoma | 1.24E-03 | 4 |
| Undifferentiated lung carcinoma | 1.35E-03 | 13 |
| Recurrent EGFR sensitizing mutation positive non-small cell lung cancer | 1.92E-03 | 4 |
| Upper airway cancer | 2.18E-03 | 66 |
| Persistent nasopharyngeal cancer | 2.35E-03 | 4 |
| Unresectable EGFR sensitizing mutation positive non-small cell lung carcinoma | 2.35E-03 | 4 |
| Sinus disorder | 2.47E-03 | 10 |
| Metastatic squamous cell cancer of the ethmoid sinus | 2.84E-03 | 4 |
| Metastatic squamous cell cancer of the glottis | 2.84E-03 | 4 |
| Metastatic squamous cell cancer of the maxillary sinus | 2.84E-03 | 4 |
| Metastatic squamous cell cancer of the supraglottis | 2.84E-03 | 4 |
| Recurrent squamous cell cancer of the ethmoid sinus | 2.84E-03 | 4 |
| Recurrent squamous cell cancer of the glottis | 2.84E-03 | 4 |
| Recurrent squamous cell cancer of the maxillary sinus | 2.84E-03 | 4 |
| Recurrent squamous cell cancer of the supraglottis | 2.84E-03 | 4 |
| Unresectable squamous cell cancer of the ethmoid sinus | 2.84E-03 | 4 |
| Unresectable squamous cell cancer of the glottis | 2.84E-03 | 4 |
| Unresectable squamous cell cancer of the maxillary sinus | 2.84E-03 | 4 |
| Unresectable squamous cell cancer of the oropharynx | 2.84E-03 | 4 |
| Unresectable squamous cell cancer of the supraglottis | 2.84E-03 | 4 |
| Stage IV ALK translocation positive non-small cell lung cancer | 3.20E-03 | 5 |
| Stage IB primary non-small cell lung cancer | 3.39E-03 | 4 |
| Stage II primary non-small cell lung carcinoma | 3.39E-03 | 4 |
| Unresectable squamous cell cancer of the hypopharynx | 3.39E-03 | 4 |
| Lung squamous cell carcinoma | 3.49E-03 | 55 |
| Locally advanced nasopharyngeal carcinoma | 4.01E-03 | 4 |
| Non-keratinizing nasopharyngeal carcinoma | 4.01E-03 | 4 |
| Stage IIIA primary non-small cell lung carcinoma | 4.01E-03 | 4 |
| Primary non-small cell lung carcinoma | 4.09E-03 | 5 |
| Unresectable recurrent non-small cell lung cancer | 4.62E-03 | 6 |
| Primary non-squamous non-small cell lung carcinoma | 4.70E-03 | 4 |

Supplementary Table 14. Downstream functional effects analysis results from genes identified as part of the differential gene expression and methylation correlation analysis.

| Functions Annotation | P-value | Genes |
| --- | --- | --- |
| Microtubule dynamics | 5.54E-10 | 47 |
| Formation of cellular protrusions | 1.19E-09 | 38 |
| Formation of cilia | 2.74E-09 | 23 |
| Organization of cytoskeleton | 3.01E-07 | 52 |
| Organization of cytoplasm | 1.01E-05 | 54 |
| Formation of axonemes | 4.48E-05 | 4 |
| Shape change of endothelial cell lines | 6.78E-05 | 5 |
| Cell movement | 1.06E-04 | 78 |
| Metabolism of reactive oxygen species | 1.68E-04 | 20 |
| Synthesis of reactive oxygen species | 2.53E-04 | 18 |
| Generation of reactive oxygen species | 4.22E-04 | 9 |
| Metabolism of superoxide | 5.57E-04 | 4 |
| Stimulation of cells | 6.04E-04 | 10 |
| Transport of sugar acid | 7.80E-04 | 3 |
| Uptake of ascorbic acid | 8.41E-04 | 2 |
| Transmembrane transport of ascorbic acid | 8.41E-04 | 2 |
| Migration of cd56+ natural killer cells | 8.41E-04 | 2 |
| Quantity of cytoneme | 8.41E-04 | 2 |
| Binding of muscle cells | 1.24E-03 | 4 |
| Stimulation of leukocytes | 1.60E-03 | 8 |
| Targeting of vesicles | 1.79E-03 | 3 |
| Migration of cells | 1.93E-03 | 66 |
| Calcium homeostasis of endoplasmic reticulum | 2.51E-03 | 3 |
| Apoptosis of epithelial cells | 2.69E-03 | 9 |
| Degranulation of granulocytes | 2.84E-03 | 4 |
| Movement of cilia | 2.84E-03 | 4 |
| Cell death of epithelial cells | 3.56E-03 | 12 |
| Stimulation of mononuclear leukocytes | 3.81E-03 | 7 |
| Cell spreading | 3.96E-03 | 9 |
| Assembly of non-motile cilium | 4.40E-03 | 3 |
| Adhesion of muscle cells | 4.40E-03 | 3 |
| Binding of AP1 consensus site | 4.70E-03 | 4 |

